# Integrated miRNA/cytokine/chemokine profiling reveals immunopathological step changes associated with COVID-19 severity

**DOI:** 10.1101/2021.08.04.21261471

**Authors:** Julie C. Wilson, David Kealy, Sally R. James, Katherine Newling, Chris Jagger, Kara Filbey, Elizabeth R. Mann, Joanne E. Konkel, Madhvi Menon, Sean B. Knight, Angela Simpson, CIRCO collaborative group, John R. Grainger, Tracy Hussell, Paul M. Kaye, Nathalie Signoret, Dimitris Lagos

## Abstract

Circulating microRNAs (miRNAs) are exceptional mechanism-based correlates of disease, yet their potential remains largely untapped in COVID-19. Here, we determined circulating miRNA and cytokine and chemokine (CC) profiles in 171 blood plasma samples from 58 hospitalised COVID-19 patients. Thirty-two miRNAs were differentially expressed in severe cases when compared to moderate and mild cases. These miRNAs and their predicted targets reflected key COVID-19 features including cell death and hypoxia. Compared to mild cases, moderate and severe cases were characterised by a global decrease in circulating miRNA levels. Partial least squares regression using miRNA and CC measurements allowed for discrimination of severe cases with greater accuracy (87%) than using miRNA or CC levels alone. Correlation analysis revealed severity group-specific associations between CC and miRNA levels. Importantly, the miRNAs that correlated with IL6 and CXCL10, two cardinal COVID-19-associated cytokines, were distinct between severity groups, providing a novel qualitative way to stratify patients with similar levels of proinflammatory cytokines but different disease severity. Integration of miRNA and CC levels with clinical parameters revealed severity-specific signatures associated with clinical hallmarks of COVID-19. Our study highlights the existence of severity-specific circulating CC/miRNA networks, providing insight into COVID-19 pathogenesis and a novel approach for monitoring COVID-19 progression.

## INTRODUCTION

The COVID-19 pandemic has caused more than 4.1 million deaths (as of 21^st^ July 2021) (1). More than 191 million people have been infected with SARS-CoV-2, the single-stranded RNA betacoronavirus that causes the disease. Infection results in a broad range of outcomes, from asymptomatic infection to lethal pneumonia, acute respiratory distress syndrome, and immunothrombosis (2, 3). Significant disease heterogeneity is observed even within hospitalised individuals with COVID-19, with some requiring only management within a ward environment and modest supplemental oxygen support while others develop critical disease requiring invasive ventilation in intensive care units (ICUs). Age, comorbidities, host genetics, and the type of immune response are all thought to play a decisive role in shaping the trajectory of the infection (4).

There is a precarious balance between protective immunity and immune-driven pathology in COVID-19, with numerous studies identifying systemic and tissue correlates of severe and fatal disease. Systemic clinical and immune correlates of severity include elevated CRP and D-dimer levels, increased numbers of neutrophils, lymphopenia, T cell exhaustion, increased number of proliferating plasmablasts and extrafollicular B cell activation, and generation of platelet-sequestering non-classical monocytes (2, 5-10). Consistent with the acute respiratory distress syndrome (ARDS) presentation of severe COVID-19, multiple studies have also reported increased circulating levels of pro-inflammatory cytokines and chemokines associated with COVID-19 severity including IL6, IL10, CCL2, CXCL10, and GMCSF (11-16). Other cytokines and chemokines, have also been reported to predict severity, although less consistently (11, 12, 15-18).

Circulating microRNAs (miRNAs) have been shown to be excellent disease diagnostic or prognostic biomarkers in a wide range of chronic and acute inflammatory and infectious diseases including viral respiratory infection. Crucially, circulating miRNA levels are thought to reflect the state of the diseased tissue (19-22). Despite their proven value as mechanism-based clinical stratification indicators, miRNAs have only minimally been explored in the context of COVID-19. Initial studies have identified potential changes in miRNA expression in COVID-19 and signatures from targeted testing of 41 miRNAs have been linked to ICU treatment and mortality in a Spanish cohort (23). Correlations between specific miRNAs and response to treatment (24) or D-dimer levels and risk of thrombosis in COVID-19 patients have also been proposed (25).

Here, we performed cytokine, chemokine and miRNA profiling in 171 blood plasma samples from 58 hospitalised patients with mild, moderate, or severe COVID-19. We found that integrating clinical parameters with systemic correlates of immune response and tissue status (cytokines, chemokines, and miRNAs) revealed qualitative changes occurring between clinical severity stages in COVID-19 and novel correlates of severe COVID-19. These severity group-specific qualitative features of COVID-19 could only be observed by combining miRNA and cytokine/chemokine profiling. Importantly, cytokine and chemokine abundance and clinically measured parameters were linked to disease severity-specific miRNA subsets, supporting that COVID-19 shows a step-wise progression in hospitalised individuals. Our findings provide novel insight into trajectories underpinning severe disease in symptomatic COVID-19 patients and support that integrating miRNA and cytokine/chemokine profiling can shed light into the severity-dependent relevance of haematological and immunopathological features of COVID-19.

## RESULTS

### Severe COVID-19 is linked to distinct plasma miRNA signatures

We performed circulating miRNA profiling as a nucleic-acid-based approach to discovering peripheral correlates of disease in COVID-19 (**Supplemental Figure 1A**). We analysed samples from 58 hospitalised patients recruited between April and November 2020 in four hospitals in Manchester, UK as part of the CIRCO cohort (**Supplemental Table 1** – cohort characteristics) (13). Patients were categorised in three groups of disease severity, mild (N=20), moderate (N=21), and severe (N=17), based on the requirement for respiratory support as described before (13). Clinical measurements and comorbidity and symptom data were also available (**Supplemental Tables 2, 3**). For several patients, follow-up plasma samples were analysed, collected at different time-points (typically taken every 1-2 days, **Supplemental Table 4**).

Expression of miRNAs was determined by Nanostring profiling and 280 miRNAs were detected in at least 10% of the samples (see Materials and Methods, **Supplemental Tables 5-6**). First, we identified miRNAs that were differentially expressed in samples from individuals with severe disease vs the rest of the cohort (moderate and mild disease). When all samples were included in the analysis (N=171 samples) we identified 32 significantly differentially expressed (DE) miRNAs (False Discovery Rate, FDR<0.1; **Figure 1A** and **Supplemental Table 5**). Down-regulated miRNAs in severe disease samples (25 in total) included hsa-miR-323a-3p, hsa-miR-451a, hsa-miR-188-5p, and hsa-miR-432-5p, while up-regulated miRNAs (8 in total) included hsa-miR-4454+7975, hsa-miR-let-7a-5p, hsa-miR-let-7f-5p, hsa-miR-630, hsa-miR-24-3p, and hsa-miR-29b-3p (**Figure 1B, Supplemental Table 5**). We repeated the analysis including only the first available sample for each patient (N=58 samples). Although, differences in miRNA expression did not reach significance after correcting for multiple testing, we identified six miRNAs which were DE in severe samples at p<0.05 and absolute log_2_ fold change (absLF)>1) (**Figure 1C, D** and **Supplemental Table 5**). These were hsa-miR-4454+7975, hsa-miR-1285-5p, hsa-miR-320e, and hsa-miR-29b-3p, which were all up-regulated, and hsa-miR-451a and hsa-miR-144-3p, which were down-regulated. hsa-miR-4454+7975, hsa-miR-29b-3p and hsa-miR-451a were identified in both analyses (including all samples or first samples). For these DE miRNAs we did not observe any significant changes in expression over the duration of hospitalisation (not shown). Having determined miRNA signatures associated with severe COVID-19 in hospitalised individuals, we performed functional association analysis using the TAM 2.0 tool (26). Using the list of DE expressed miRNAs in severe cases (corresponding to **Figure 1A**), enriched functional terms for up-regulated miRNAs included cell death, innate immunity, and cell cycle (**Figure 1E**). For down-regulated DE miRNAs, enriched terms included Regulation of Stem Cell, Vascular inflammation, and Chemotaxis (**Figure 1E**). This indicated terms that were likely to reflect the immune response to infection (e.g. Innate immunity) but also terms that could reflect tissue pathology (e.g. cell death). This was also the case, when performing functional term enrichment analysis using the list of all DE miRNAs based on first available samples only (corresponding to **Figure 1B**), with terms referring to wound healing and extracellular matrix remodeling being most enriched (**Supplemental Figure 1B**). We note that statistical significance was lower due to the smaller number of DE miRNAs. We then explored functional associations of top predicted targets of up-regulated miRNAs in severe samples using the Targetscan tool (27) (**Supplemental Figure 1C**). Notably enriched gene ontology process associations included terms involving extracellular matrix remodeling and responses to hypoxia consistent with severe COVID-19 pathology (**Figure 1F**). Overall, our analyses revealed a miRNA signature associated with severe COVID-19 and suggested that this signature could be reflective of tissue pathology and damage.

**Figure 1:**
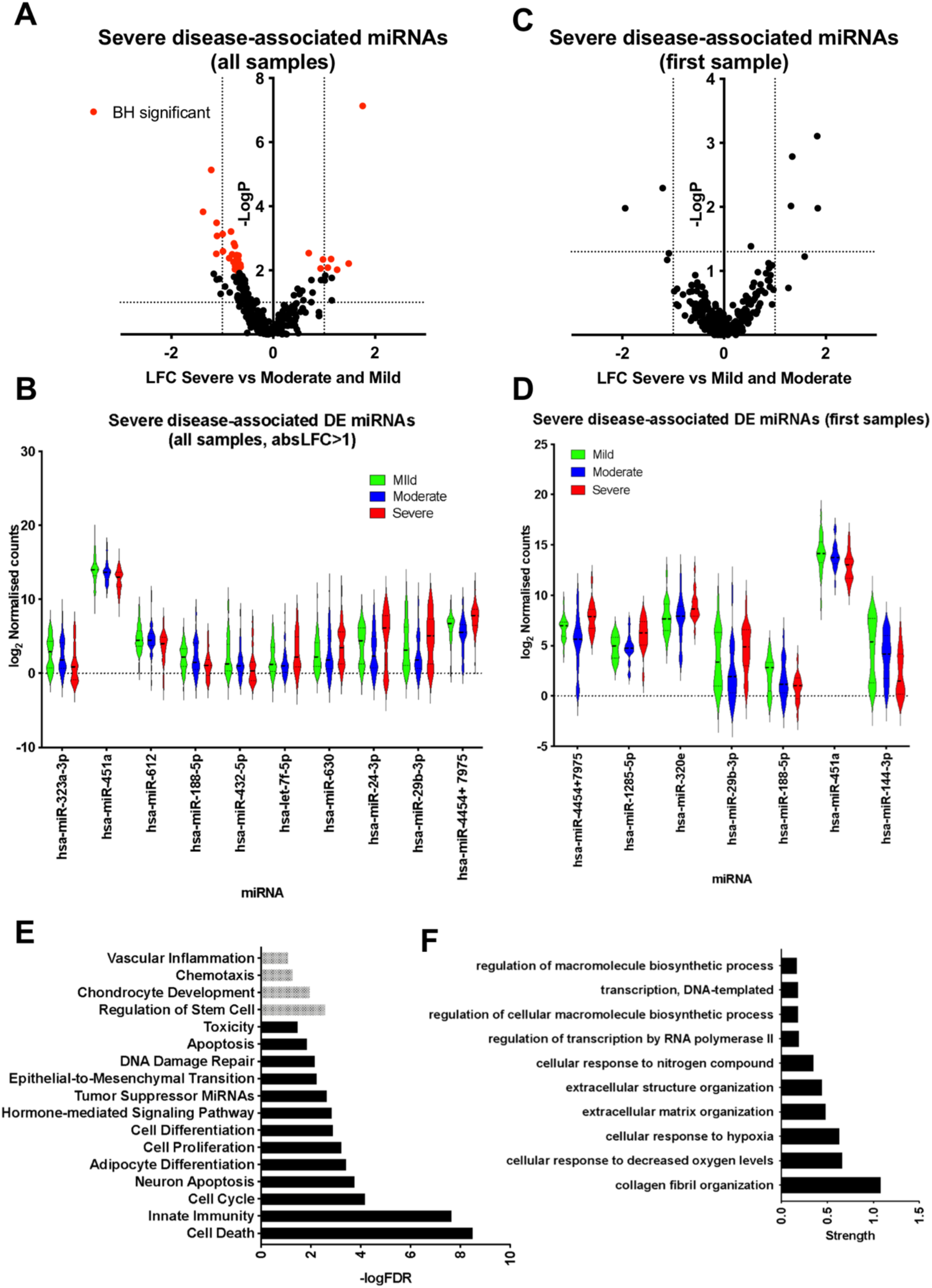
Plasma miRNA signatures of severe COVID-19. **A**. Volcano plot comparing miRNA expression in severe against mild and moderate samples including all samples as independent observations. Dotted lines correspond to log_2_ fold change (LFC) greater than one and Mann Whitney p < 0.05. Red dots correspond to miRNAs for which FDR<0.1 following Benjamini Hochberg (BH) correction. **B**. Violin plots of top DE expressed miRNAs in severe samples (FDR < 0.1 and absLFC>1), including all samples. **C**. Volcano plot comparing miRNA expression in severe against mild and moderate samples including only the first available samples for each patient. Dotted lines correspond to log_2_ fold change (LFC) greater than one and Mann Whitney p < 0.05. **D**. Violin plots of DE expressed miRNAs in severe samples when only the first sample per patient is included (p < 0.05 and absLFC > 1). **E**. Significantly over-represented functional terms within up-regulated (shown in black) and down-regulated (shown in grey) miRNAs in severe cases (shown in red in Figure 1A), including all samples as independent variables. **F**. Significantly over-represented gene ontology process terms amongst predicted targets of miRNAs up-regulated in severe cases (red dots in the top right quadrant in Figure 1A).

### Progression from mild to moderate and severe COVID-19 is characterised by a global reduction in circulating miRNA levels

Next, we explored miRNA signatures of mild COVID-19 disease within hospitalised individuals. When performing comparisons between individuals with mild disease vs the rest of the cohort (moderate and severe disease), either including all samples or just first available samples, the most notable finding was a striking global (for the vast majority of detectable miRNAs) decrease in circulating miRNA levels in individuals with severe and moderate disease when compared to individuals with mild disease (**Figure 2A-D**). When including all samples, mild cases were characterised by significantly higher levels (FDR<0.1) of miRNAs including hsa-miR-495-3p, hsa-miR-150-5p, hsa-miR-432-5p, hsa-miR-376a-3p, hsa-miR-146a-5p, and hsa-miR-451a (**Figure 2B, Supplemental Table 6**). When including only the first available sample for each patient differences were, as in the case of severe disease, more modest (p<0.05, absLFC>1), and DE miRNAs incldued hsa-miR-548g-3p, hsa-miR-150-5p, hsa-miR-590-5p, hsa-miR-518b, hsa-miR-363-3p, and hsa-miR-495-3p (**Figure 2C-D, Supplemental Table 6**). Functional association analysis, using TAM 2.0, indicated an enrichment in miRNAs contributing to immune processes amongst DE miRNAs in mild COVID-19 (corresponding to **Figure 2A**), and more generic cellular processes when considering DE miRNAs only in first samples (**Figure 2E**). Notably, exploring gene ontology terms amongst the top predicted targets of up-regulated miRNAs (**Figure 2A**), we observed enrichment in functional terms associated with mRNA and small RNA metabolism (**Figure 2F**). This could reflect the type response against SARS-CoV-2 and can be directly linked to the higher circulating miRNA levels in patients with mild disease. These analyses revealed a miRNA signature associated with mild COVID-19, reflecting elements of both the immune response to SARS-CoV-2 infection but also tissue pathology.

**Figure 2:**
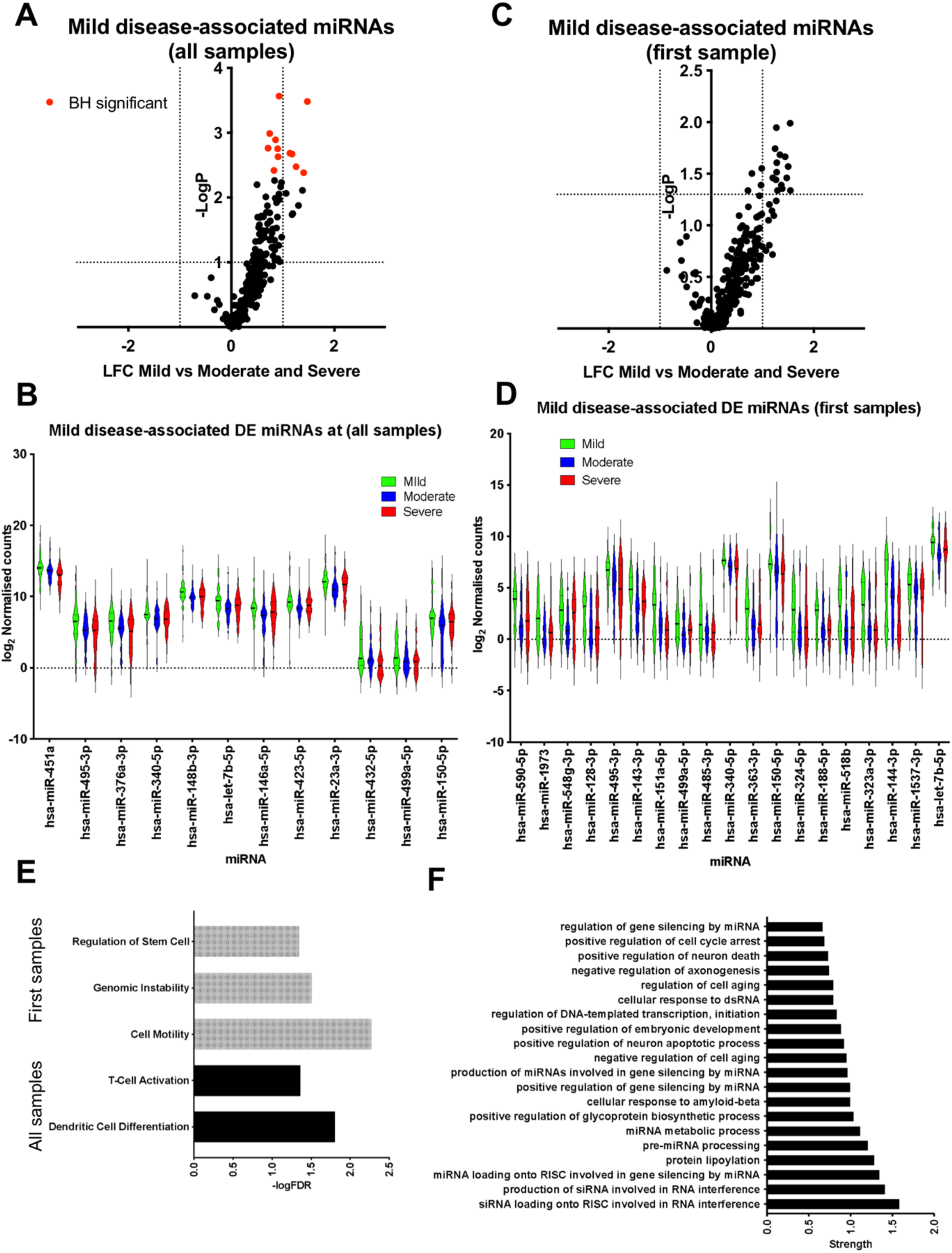
Plasma miRNA signatures of mild COVID-19. **A**. Volcano plot comparing miRNA expression in mild against moderate and severe samples including all samples as independent observations. Dotted lines correspond to log_2_ fold change (LFC) greater than one and Mann Whitney p < 0.05. Red dots correspond to miRNAs for which FDR<0.1 following BH correction. **B**. Violin plots of DE expressed miRNAs in mild samples (FDR < 0.1), including all samples. **C**. Volcano plot comparing miRNA expression in mild against moderate and severe samples including only the first available samples for each patient. Dotted lines correspond to log_2_ fold change (LFC) greater than one and Mann Whitney p < 0.05. **D**. Violin plots of DE expressed miRNAs in mild samples when only the first sample per patient is included (p < 0.05 and absLFC > 1). **E**. Significantly over-represented functional terms for miRNAs up-regulated in mild samples when all samples (shown in black) or only first samples (shown in grey) are analysed. **F**. Significantly over-represented gene ontology process terms amongst predicted targets of miRNAs up-regulated in mild cases (red dots in the top right quadrant in Figure 2A).

### Cytokine and chemokine profiling reveal quantitative and qualitative differences between COVID-19 severity groups

In addition to miRNA profiling, we measured cytokine (**Figure 3A, B**) and chemokine (**Figure 3C, D**) levels in the same plasma samples. When including all samples as independent variables we observed significant (FDR<0.05) up-regulation of most tested cytokines in severe samples (**Figure 3A, Supplemental Table 7**). However, when taking only the first available samples for each patient only up-regulation of IL-6 reached statistical significance when comparing severe samples to moderate and mild (**Figure 3B**). With regards to circulating chemokine levels, CCL3, CXCL9, CCL20, and CXCL1 were significantly up-regulated in samples from individuals with severe COVID-19 when compared against mild and moderate cases (**Figure 3C**). Repeating the analysis to only include first samples, showed again increased levels of CXCL9 and CCL20 in severe disease, but also increased levels of CXCL10 and a down-regulation of CCL17 in patients with severe COVID-19 (**Figure 3D**). Comparison of mild samples against the rest of the cohort (moderate and severe) revealed significantly lower levels of CXCL9, CCL20, IL6, IL10, IFNγ, and IL4 in mild samples, whereas levels of CCL5 were significantly increased (**Supplemental Table 7**). Within the studied sampling time-frame we did not observe any statistically significant patterns of regulation for DE cytokines and chemokines over time (not shown).

**Figure 3:**
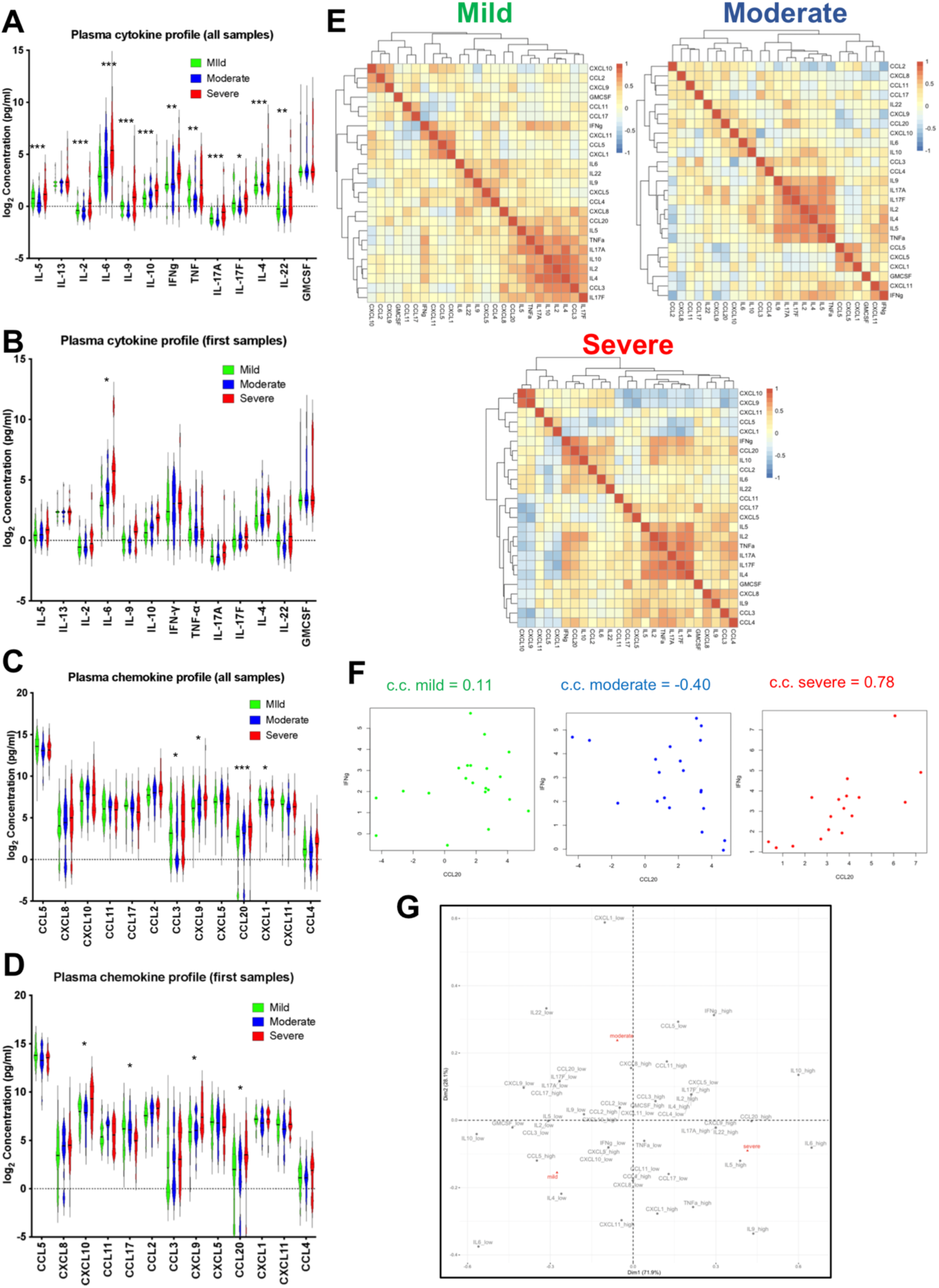
Circulating cytokine and chemokine signatures of COVID-19. **A**. Violin plots of measured cytokines in mild (green), moderate (blue), and severe (red) samples, including all samples. Stars indicate significance for the comparison of severe against mild and moderate as follows: * FDR < 0.05, ** FDR < 0.01, *** FDR < 0.005. **B**. As in A, but analysis including only first available samples per patient **C**. As in A, but for circulating chemokines **D**. As in C, but analysis including only first available samples per patient **E**. Heatmaps showing Spearman correlation between cytokines and chemokines for mild, moderate, and severe groups separately. The average value over all available measurements for each patient was used in the calculation of correlations. **F**. Plots of IFNγ against CCL20 levels in mild (green), moderate (blue), and severe (red) patients. Correlation coefficients (c.c.) are shown above each scatter plot. **G**. Correspondence analysis for cytokines and chemokines showing the association between severity groups and high and low values of cytokines and chemokines.

We performed correlation analysis between individual cytokines and chemokines, taking each sample as an independent observation (**Supplemental Figure 2A**) or only including the first available sample for each patient (**Supplemental Figure 2B**). However, we also performed analyses using the average levels for each patient. This approach has the benefit of both capturing a representative and stable cytokine profile per patient (by including multiple sampling points for most patients, **Supplemental Table 4**) and avoiding having over-representation of some patients due to a higher number of collected samples. The above-mentioned lack of significant changes in cytokine and chemokine levels over the studied period meant that by taking an average of all measurements we were less likely to misrepresent the levels of any specific protein in the correlation matrix. Following this approach, we found that IL2, IL4, IL17A, IL17F and TNF were positively correlated in all three severity groups (**Figure 3E, Supplemental Figure 2C**), as were IL5 and IFNγ but to a lesser extent particularly in the moderate group (**Figure 3E, Supplemental Figure 2A, B**). Notably, IL10 was also part of this correlation cluster but only for patients with mild disease. Furthermore, CCL5 and CXCL1 were positively correlated in all groups. CXCL11 and CXCL5 were positively correlated with these, but only in mild and moderate disease groups respectively. CXCL10 and CXCL9 also correlated in all groups, although the correlation was stronger in severe cases (**Figure 3E, Supplemental Figure 2D**). IL9 was positively correlated with CCL3, CCL4, and CXCL8 in severe samples but to a lesser extent, or not at all in moderate and mild samples, respectively (**Figure 3E**). CXCL1 and IL17F were negatively correlated only in moderate and severe samples, the effect being stronger in severe. The same was observed for CXCL10 and CCL17. Notably, we found a drastic qualitative difference between the three severity groups with regards to correlation between CCL20 and IFNγ, which were not correlated in mild, negatively correlated in moderate, and positively correlated in severe (**Figure 3F**). In addition, to identify relationships between severity groups and high or low levels of cytokines or chemokines we preformed correspondence analysis with count data obtained by assigning values as low, medium or high for each severity group (see Methods). This showed that, while most high values are associated with the severe group, high values of CCL5 are associated with mild samples (**Figure 3G**). Furthermore, correspondence analysis indicated that IL6 (p < 0.001), IL9 (p < 0.05), IL10 (p < 0.01), and CXCL1 (p < 0.05) were significantly associated with severity. Overall, the cytokine and chemokine profiling presented here and correlation analyses per severity group demonstrated that in addition to quantitative, there were also qualitative changes between severity groups.

### Integration of miRNA and cytokine/chemokine profiles reveals severity group-specific circulating molecular signatures of COVID-19

To gain more insight into potential step changes between severity groups we performed partial least squares regression (PLSR) analysis on the miRNA and cytokine and chemokine profiles together (including all samples as independent observations). We found separation between samples from patients with severe disease and those with mild or moderate disease (**Figure 4A** and **Supplemental Figure 3A**). Using PLS models, we performed leave-one-patient-out (L-O-P-O) classification to assess the ability to predict severity based on our dataset. As discrimination between mild and moderate samples was poor, we repeated the analysis with just two classes, severe vs mild/moderate. The analysis was performed using only cytokine and chemokine data, all miRNAs, only DE miRNAs, and cytokine and chemokine data together with DE miRNAs. We found that combining the cytokine/chemokine levels with DE miRNAs resulted in the highest overall accuracy (87%) in severity classification (**Figure 4B**). To prevent individual patients with multiple samples dominating the analysis, we calculated correlations using average values (one per patient). Notably, correlations between miRNAs and cytokines/chemokines calculated across the whole cohort were generally weak (**Supplemental Figure 3B**). However, when performing the analysis within severity groups stronger and distinct correlations were discovered (**Supplemental Figure 3C** and **Supplemental Table 8**). The number of these correlations was notably higher in the severe disease group (**Supplemental Figure 3C**, miRNAs and cytokines/chemokines that show at least one significant correlation with correlation coefficient higher than 0.4). We observed severity group-specific features. CXCL1 showed positive correlations with a high number of miRNAs in severe disease compared to mild and moderate. CXCL1 also clustered with CXCL9 and CXCL10 in severe disease but not in moderate. IL6 showed several positive correlations with miRNAs and clustered with IL10, CCL20, GMCSF, IL22, and IFNγ in severe samples, and this was not the case for moderate and mild groups (with the exemption of CCL20). Interestingly, IL17F showed predominantly negative correlations with miRNAs again in manner specific to the severe disease group (**Supplemental Figure 3C**). Strong, severity-group specific correlations between CCL17 and miR-92a-3p (mild), CXCL1 and miR-1283 (moderate), IL-17F and miR-425-5p (severe), and CXCL9 and miR-1246 (severe) were also observed (**Figure 4B**). It is of note that, although these correlations were severity group-specific, none of these miRNAs were significantly differentially expressed in severe or mild samples (**Supplemental Tables 5-6)**.

**Figure 4:**
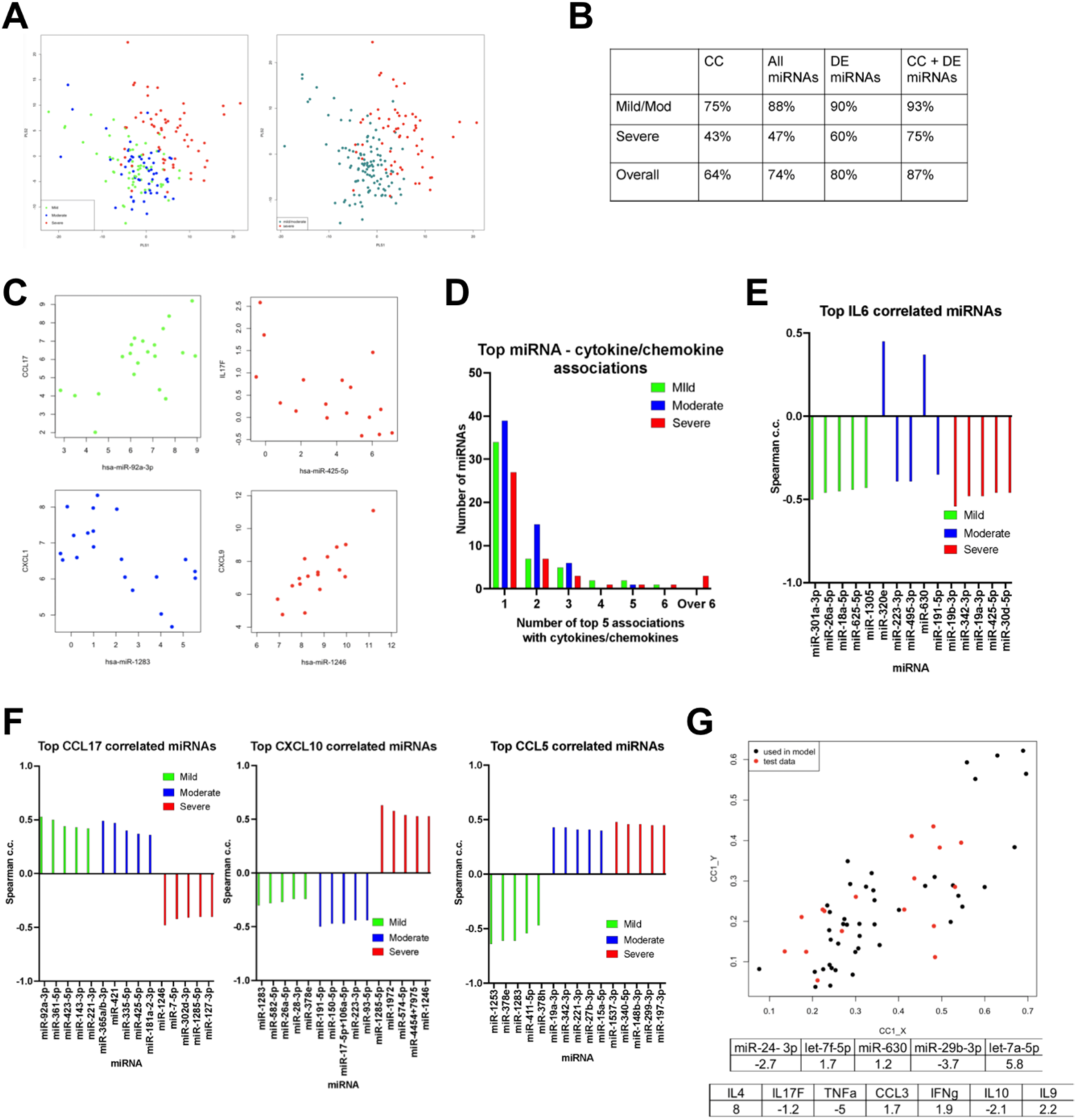
Integration of miRNA, cytokine, and chemokine signatures. **A**. Score plots for the first two latent variables from PLSR obtained using cytokine and chemokine data together with DE miRNA measurements. The plot on the left was obtained using all three severity levels (mild, moderate, and severe) whilst that on the right was obtained using only two classes (severe and mild/moderate). **B**. Results from PLSR with patients considered in two groups for different datasets with continuous output values assigned to the nearest discrete class number. Accuracies shown were obtained using leave-one-patient-out cross validation, using only cytokine and chemokine (CC) values, all miRNA values, only DE miRNAs values, or combining all CC values with DE miRNAs. **C**. Example scatter plots showing strong, severity group-specific correlations in mild (green), moderate (blue), or severe (red) patients. **D**. Number of miRNAs (Y-axis) in the top 5 strongest correlations with 1, 2, 3, 4, 5, 6, or over 6 cytokines/chemokines per severity group. See Supplemental Table 4. **E**. Spearman correlation coefficients for top 5 correlated miRNAs with IL6 in mild (green), moderate (blue), and severe (red) groups. **F**. As in E, but for CCL17, CXCL10, and CCL5. **G**. Canonical correlation analysis (CCA) to relate the miRNAs associated with Cell Death in the severe group (Figure 1E) with cytokines and chemokines. Cytokines/chemokines with individual absolute correlation greater than 0.4 (for the severe group) with any of these miRNAs were included in the analysis. The canonical scores plot shows data (from individual samples) for the12 patients used to determine the canonical covariates (c.c. = 0.8, p = 4.8 × 10^−6^) in black. The coefficients obtained are shown in the table and are used to obtain the canonical scores for 5 randomly chosen patients reserved as an independent test set (plotted in red).

### Distinct miRNA correlates of cytokine/chemokine expression underpin severe COVID-19

For differentially expressed cytokines and chemokines in severe or mild samples we determined the five miRNAs with the highest absolute correlation coefficients per severity group (**Supplemental Table 8**). In addition to severity group-specific top miRNA correlates of plasma cytokine and chemokine levels, this analysis revealed a striking qualitative difference for the severe group, namely the increased number of miRNAs featuring as top correlating with multiple cytokines and chemokines (**Figure 4D**). In severe patients, miR-30d-5p was in the top 5 most correlated miRNAs for 9 cytokines and chemokines. Furthermore, hsa-miR-425-5p and hsa-miR-19a-3p feature as top correlates of 8 cytokines and chemokines, with hsa-miR-19b-3p featuring a further 6 times (**Supplemental Table 8**). There were no miRNAs in the top 5 lists of more than 6 cytokines and chemokines in mild or moderate cases. Conversely just 27 miRNAs appeared only once as one of the top 5 strongest correlations in severe cases, the number increasing to 39 and 34 for moderate and mild cases, respectively (**Figure 4D**). Of note, we observed distinct top correlating miRNA for each severity group for all tested cytokines and chemokines, including for example IL6, a cytokine widely associated with COVID-19 severity (**Figure 4E**). In addition to different miRNAs, we found several cytokines with opposing correlation trends. For example, all top strongest miRNA correlations for CCL17 were negative in the severe group but positive in the mild and moderate, the reverse being the case for CXCL10. Similarly, the mild group was characterised by all top CCL5 correlations being negative in contrast to moderate and severe groups (**Figure 4F**).

Interestingly, the above cross-correlation analysis (**Supplemental Figure 3C**) with miRNA expression separated in a severity-dependent manner the IL2, IL4, IL17A, IL17F, TNF cluster found in **Figure 3E** (when only cytokine and chemokine expression were considered). This suggested that co-expression of these pro-inflammatory cytokines might be associated with distinct immunological and/or tissue pathology effects in each subgroup. We further explored this by focusing on miRNAs that contributed to enrichment of the Cell Death functional association amongst DE miRNAs in severe samples (**Figure 1E**). These miRNAs were miR-24-3p, Let-7f-5p, miR-630, miR-29b-3p, and Let-7a-5p. We reasoned that a set of chemokines or cytokines that would correlate with these miRNAs as a group could potentially be primary candidates to be effectors of lung tissue damage. We used canonical correlation analysis (CCA) to relate a linear combination of the miRNAs associated with cell death with a linear combination of the cytokines and chemokines that showed the highest correlations individually with at least one of these miRNAs in severe samples (**Supplemental Table 9**). The coefficients in the linear combinations with maximal correlation, obtained using samples from only 12 of the 17 patients in the severe group, are given in **Figure 4G**. For this data, the correlation for this first canonical pair was 0.8 with a p-value of 4.79e-06, identifying IL4 and TNF as the predominant contributors to the correlation with Cell Death miRNAs, with minor contributions from IL17F, CCL3, IFNγ, IL9, and IL10. We note here that collinearity due to correlations within these cytokines and chemokines may make individual contributions difficult to estimate. Applying the coefficients to the data for samples from the remaining 5 severe patients showed that the model generalises to unseen data with the correlation over all severe samples being 0.76. The canonical variates are plotted for both training and test data in **Figure 4G**.

### Integration of miRNA and cytokine/chemokine profiles with clinical parameters in hospitalised COVID-19 patients

The above findings identified systemic signatures that were both quantitatively and qualitatively different between COVID-19 severity groups. We therefore explored how these could potentially underpin clinical features of COVID-19 by correlating levels of cytokines and chemokines with clinical measurements. For this analysis we considered cytokine and chemokine levels from the first available plasma samples as this was the timepoint matching the timepoint of acquisition of clinical measurements.

In severe samples, we found a cluster including IL6, IFNγ, CCL2, and CCL20 positively correlating with ALT, CRP, saturated O_2_, and creatinine and negatively correlating with HCT (**Figure 5A**). A second cluster including TNF, IL17A, IL17F, IL9, IL2, and IL4 correlated positively with several cell populations including neutrophils and eosinophils but largely negatively with monocytes and lymphocytes. This clustering profile was specific to severe cases. Several strong and group-specific correlations were observed. IL9 correlated positively and strongly with lymphocytes in moderate cases, modestly in mild, and not at all in severe (**Figure 5A**). Other group-specific correlations included IL17F with BMI and CCL11 with temperature in mild, CCL11 with MCV and CXCL10 with WCC in moderate, and CCL5 with potassium and CXCL9 with ALP in severe (**Figure 5B**). Interestingly, we noted that cytokines widely associated with COVID-19 severity such IL6, CXCL10, and TNF showed multiple but modest correlations with clinical parameters in all groups, likely reflecting their multifunctional role in disease development.

**Figure 5:**
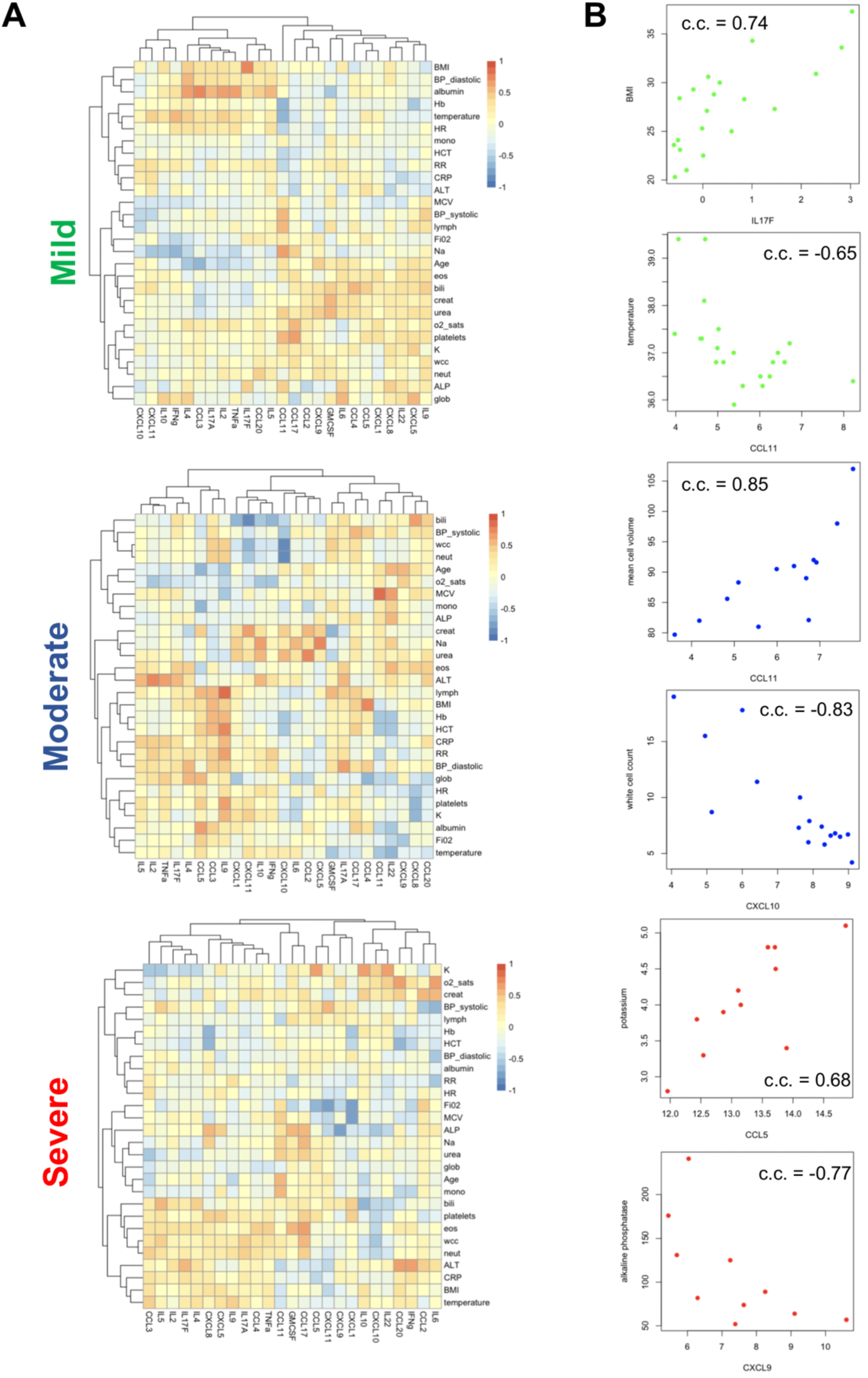
Correlation analysis of cytokine, and chemokine profiles with clinical parameters. **A**. Heatmaps showing Spearman correlation coefficients (c.c.) between cytokines and chemokines and clinical measurements for mild, moderate, and severe groups. The first cytokine/chemokine value was used for each patient. **B**. Example scatter plots for strong, severity group-specific correlations between cytokines/chemokines and clinical parameters in mild (green), moderate (blue), or severe (red) patients. Correlation coefficients (c.c.) also shown.

We also investigated significant correlations between levels of individual miRNAs (often not significantly DE in severe samples when compared to moderate and mild) and clinical measurements. The clinical parameters separated in different clusters between groups and the number of strong correlations (absolute value > 0.4) was higher in moderate cases (**Figure 6A-C**). Of note clinical parameters, previously associated with COVID-19 severity, separated in different clusters between groups. For example, albumin and globulin (28) clustered together in mild and moderate samples but not in severe (**Figure 6A-C**) and the top miRNA correlates of albumin and globulin were distinct between severity groups (**Figure 6D**). In addition, the top miRNA correlates of lymphocyte numbers showed positive correlations in the moderate and severe groups but negative correlations in the mild, whilst we also observed severity group-specific correlations between miRNAs and neutrophils (**Figure 6D**). Interestingly, for CRP (**Figure 6D**) and haemoglogin (**Supplemental Figure 4**), we found switches in the type of correlation and identity of miRNAs between mild and moderate cases, whereas only modest correlations were observed in the severe group (**Figure 6C**). Similar group-specific correlations were observed for all tested clinical parameters (**Supplemental Figure 4**). In all groups we observed multiple positive correlations between platelets and individual miRNAs (**Figure 6E**). However, we did not find any miRNAs that correlated with platelets in all groups. Eleven miRNAs strongly correlated with platelets in at least two groups including miR-93-5p, miR-361-5p, miR-423-5p, miR-106a-5p/miR-17-5p in mild and moderate, miR-376c-3p in moderate and severe, and miR-382-5p, miR-503-5p, miR-590-5p, miR-652-3p, miR-151a-3p, and miR-323a-3p in mild and severe. Overall, integration of cytokine, chemokine, and miRNA profiles with clinical parameters identified commonalities but also significant differences between disease severity groups in hospitalised individuals.

**Figure 6:**
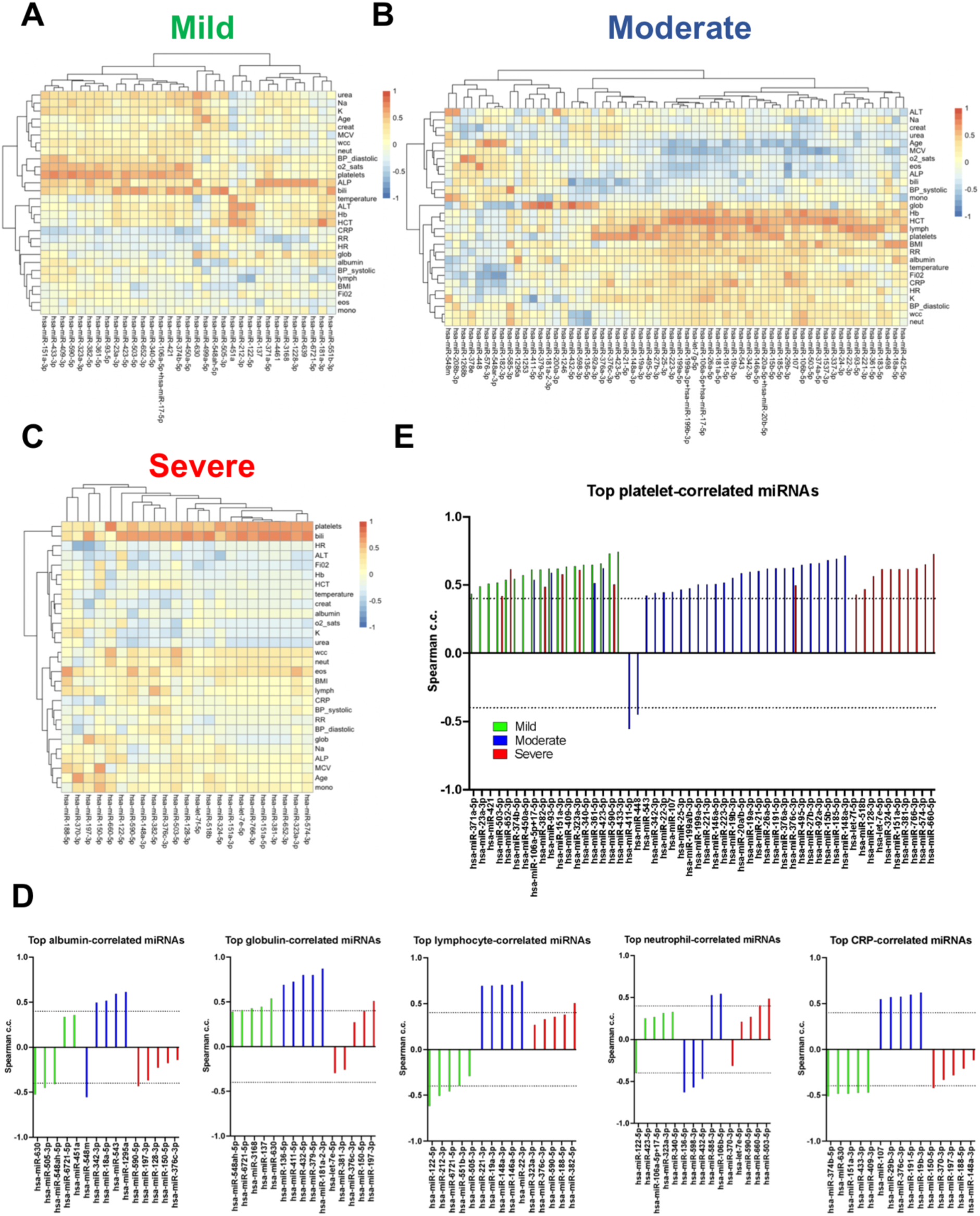
Severity-specific correlations between miRNAs and COVID-19-associated clinical parameters. **A**. Heatmap showing Spearman correlation coefficients (c.c.) between miRNAs and clinical measurements for the mild group of patients. Values shown for miRNAs with absolute c.c. > 0.6 with at least one clinical parameter. The first miRNA value was used for each patient. **B**. As in A but for moderate cases. **C**. As in A but for severe cases **D**. Spearman correlation coefficients for top 5 correlated miRNAs with indicated clinical parameters in mild (green), moderate (blue), and severe (red) groups. **E**. miRNAs with absolute c.c. > 0.4 with platelets in mild (green), moderate (blue), and severe (red) groups.

## DISCUSSSION

Our integrated profiling of circulating miRNA, cytokine, and chemokine levels and linkage to clinical parameters revealed new insights into quantitative and, crucially, qualitative changes occurring during COVID-19 progression in hospitalised patients. Our study represents the largest circulating miRNA profiling study in COVID-19 patients. Using miRNA profiling to deconvolute cytokine and chemokine abundance resulted in identifying powerful qualitative distinctions between severity groups, including sets of miRNAs that correlate in a severity-specific manner with cytokines and chemokines already associated with COVID-19 severity such as IL6 and CXCL10 (29). Although increased levels of pro-inflammatory factors are associated with severe disease, previous reports (11-16) and this study show significant overlap in their circulating levels between severity groups. Significantly, therefore, we show that by identifying correlations between miRNAs and these cytokines and chemokines we can robustly identify patients with severe disease in a manner not possible through analysis of cytokines/chemokines alone. Our approach not only reveals effects that occur only in severe COVID-19, but can also be used as a foundation for stratifying patients and understanding response to treatments such as anti-IL6R blocking antibodies (30). Indeed, small-scale studies have proposed candidate miRNA predictors of response to IL6R blockade (24). It is of note that several of the severity group-specific miRNA-cytokine or miRNA-clinical parameter correlations we report involve miRNAs that are not significantly differentially expressed in severe or mild samples. This indicates the emergence of disease-associated novel networks rather than just quantitative increase of interactions operating in a continuum during progression from mild to moderate to severe disease. Interestingly, although such qualitatively changes are more numerous and profound when combining miRNA and cytokine/chemokine profiling, a small number can be already determined when exploring only cytokine and chemokine levels. In this respect, IFNγ and CCL20 show a strong positive correlation only in patients with severe disease. This correlation and the increase in plasma concentrations seen for both markers could be related to severe lung inflammation rather than a systemic immune response. Indeed, IFNγ is an essential trigger of antimicrobial pulmonary responses, but increased IFNγ levels in COVID-19 blood and airways samples are associated with pathogenesis and pulmonary tissue damage (31-33), possibly through a detrimental effect of IFNγ on epithelial cells (34). On the other hand, CCL20 is secreted by the bronchial epithelium and important for lung immunity, but it can lead to pathogenic responses (35). COVID-19 studies have already reported increased CCL20 levels in blood as well as respiratory specimen from severe disease patients and is linked to ARDS (31, 36, 37).Therefore, the strongly correlated IFNγ and CCL20 may mark a circulating readout of severe respiratory disease complications.

We propose that circulating miRNA profiling can provide insight into effects happening at the tissue level adding a valuable methodological modality to cytokine and chemokine profiling. A correlation between circulating and diseased tissue profiles has been extensively reported in cancer, and autoimmune and infectious diseases (19-22). Our findings show that both circulating miRNAs and their predicted targets are indeed reflective of processes occurring at the tissue level (e.g. cell death, response to hypoxia, extracellular matrix remodelling). This provides a powerful tool in further understanding main activities of multifunctional cytokines and chemokines. For example, we identify a significant linkage between IL4 and TNF and cell death-associated miRNAs. We note that these associations do not imply causality, as changes in circulating cytokine and chemokine levels can also be the consequence of tissue damage. Nevertheless, they provide a potential tool that can be used as a peripheral window into tissue pathology. Interestingly, a smaller study (38), has compared 10 COVID-19 patients with 10 controls and identified 55 DE miRNAs in COVID-19 patients. Seven of those, (hsa-let-7a-5p, hsa-let-7f-5p, hsa-miR-150-5p, hsa-miR-423-5p, hsa-miR-432-5p, hsa-miR-451a, hsa-miR-651-5p) were also DE in severe or mild patients in our study, indicating an overlap between disease- and severity-associated miRNAs. As the aim of our study was to search for differences between COVID-19 severity groups in symptomatic hospitalised patients, we did not include healthy controls here.

A strength of our study was the ability to correlate with clinical laboratory measurements. We find severity group-specific associations. In severe samples, two cytokine/chemokine clusters were observed one of which (TNF, IL17A, IL17F, IL9, IL2, and IL4) correlated positively with neutrophils and largely negatively with monocytes and lymphocytes, providing some insight into inflammatory profiles associated with clinical hallmarks of severe COVID-19, namely lymphopenia and neutrophilia. Indeed, the neutrophils to lymphocytes ratio is a strong correlate of COVID-19 severity (7, 39). In parallel, miRNAs also revealed severity group-specific clusters. Of interest, several miRNAs correlate with platelets in our analysis. Although we cannot exclude the possibility that this could be partly due to platelet contamination or lysis, we note that amongst a list of 11 miRNAs previously reported to strongly correlate with platelet contamination (40) only two (hsa-miR-93-5p and hsa-miR-17-5p) correlated with platelets in more than one group in our study. Alternatively, platelet-secreted miRNAs have been reported to reflect platelet activation and function (41). In this respect, our platelet-associated miRNA signatures and the observation that they differ between severity groups provides novel insight into the observed thrombocytopenia and platelet aggregation phenotype in severe COVID-19 (42, 43).

Overall, integrated miRNA/cytokine/chemokine profiling determines key systemic changes that reflect the step-wise progression of COVID-19. Other methodologies such as whole blood RNA sequencing, single cell RNA sequencing, or autoantibody profiling also support the existence of distinct disease progression trajectories characterised by unique quantitative and qualitative features (44-48). Identifying qualitative differences between different severity manifestations of COVID-19 in symptomatic individuals and investigating their mechanistic relevance can provide important clues for development of future COVID-19 therapies. Using exclusively circulating measurements to dissect these differences means that our work already provides mechanistic clues and a set of corresponding candidate biomarkers that can be used by the clinical and research community to study pathogenesis and treatment of COVID-19. We propose that targeted tandem miRNA-cytokine/chemokine profiling provides a flexible non-invasive toolkit to help mechanistic studies, patient stratification, and response monitoring for future and current COVID-19 therapeutics.

## METHODS

### Clinical study design and sample collection

The clinical recruitment and study design has been reported elsewhere (13). Individuals with COVID-19 were recruited from 3 hospitals in the Greater Manchester area, Manchester University Foundation Trust (MFT), Salford Royal NHS Foundation Trust (SRFT) and Pennine Acute NHS Trust (PAT) under the framework of the Manchester Allergy, Respiratory and Thoracic Surgery (ManARTS) Biobank (study no M2020-88) for MFT or the Northern Care Alliance Research Collection (NCARC) tissue biobank (study no. NCA-009) for SRFT and PAT. Ethics approval was obtained from the North West-Haydock Research Ethics Committee for ManARTS (reference 15/NW/0409) and from the Wales Research Ethics Committee 4 for NCARC (reference 18/WA/0368). Participants were admitted to hospital between April and November 2020. Clinical information was extracted from written/electronic medical records. Only patients who tested positive for SARS-CoV-2 by reverse transcriptase polymerase chain reaction (RT-PCR) on nasopharyngeal/oropharyngeal swabs or sputum were included. Disease severity was scored based on degree of respiratory failure (13), as below:

Mild: less than 3L/min or less than 28% supplemental oxygen and managed in a ward. Moderate: less than 10L/min or less than 60% supplemental oxygen, managed in a ward environment; chronic NIV or CPAP (at home) or acute NIV for individuals with COPD. Severe: any of more than 10L/min or less than 60% supplemental oxygen, or use of acute NIV, or managed in ICU with invasive ventilation.

Where severity of disease changed during admission, the highest disease severity score was selected for classification. Samples were collected as soon after admission as possible and at one- or two-day intervals thereafter (**Supplemental Table 4**).

### Blood plasma isolation

Whole venous blood was collected in tubes containing EDTA (Starstedt) and diluted 1:1 with PBS. Peripheral blood mononuclear cells (PBMCs) were isolated by density gradient centrifugation using Ficoll-Paque Plus (GE Healthcare) and 50 ml SepMate tubes (STEMCELL technologies) according to the manufacturer’s protocol. Blood plasma, at the top of the gradient was collected and stored at -80°C until usage.

### RNA extraction and miRNA profiling

RNA was extracted from 800μl plasma the Norgen Plasma/Serum RNA Purification Midi kit (Norgen, Cat #56100). A spike-in non-human miRNA (osa-miR-414) was added to each sample (5μl from a 200 pM per sample) for normalisation. For miRNA profiling we used the Nanostring nCounter™ platform and the human v3 miRNA panel was used containing probes for 827 human miRNAs. Extraction of raw reads and analysis of the individual datasets was carried out using the nSolver™ Analysis Software. For each miRNA the mean plus one standard deviation of negative controls from all counts was subtracted. A minimum value of 1 to all counts. All miRNAs which were expressed above this threshold in fewer than 10% of samples were removed, leaving 280 miRNAs for further analysis. The spike-in count (osa-miR414) was used to normalise miRNA counts. Differential expression analysis on these normalised values was performed by performing Mann Whitney U test, followed by a false discovery rate (FDR) correction using the Benjamini Hochberg correction.

### Cytokine and chemokine profiling

With the exemption of GM-CSF, for cytokine and chemokine profiling bead-based multiplex flow cytometry assays were used. Cytokines and chemokines levels were quantified from patients’ plasma fractions using three sets of pre-defined LEGENDplex panels: Human Th-2 cytokine Panel version 2 (IL2, IL4, IL5, IL6, IL9, IL10, IL13, IL17A, IL17F, IL22, IFNγ and TNF), Proinflammatory chemokine Panel 1 (CCL2, CCL3, CCL4, CCL11, CCL17 and CCL20; CXCL1, CXCL5, CXCL8, CXCL9, CXCL10 and CXCL11) plus a separate CCL5 1-plex kit (BioLegend, San Diego, USA) according to manufacturer’s instructions. GM-CSF was detected and quantified using a sandwich ABTS ELISA kit (PeproTech EC, Ltd, UK). Statistical significance in differences in cytokine and chemokine expression between groups were assessed using Mann Whitney U test, followed by Benjamini Hochberg correction.

### Data integration and statistical methods

Partial least squares regression (PLSR) was performed using the R package ‘pls’ with the response variable encoded as 1.0, 2.0 and 3.0 for mild, moderate and severe observations respectively. Leave-one-patient-out (L-O-P-O) classification was carried out by assigning the class corresponding to the closest integer to the output response. For each patient, a model was built without using the data for any of that patient’s samples and the results are given for the prediction of the observations left out for each patient in turn. As results showed mild and moderate patients could not be distinguished, the L-O-P-O classification was repeated using just two classes, with mild and moderate observations combined as one class.

For correspondence analysis (CA), min-max scaling was applied to the data for each chemokine and cytokine so that each variable had a minimum of zero and maximum of 3.0 over all observations. With values less than 1.0 considered as ‘low’, values between 1.0 and 2.0 as ‘medium’ and values > 2.0 considered as ‘high’, count data for each severity group was obtained for each variable. To reduce the number of variables, medium counts were excluded from CA, which was performed using the *corresp()* function from the ‘MASS’ package in R. All count data was used in chi-squared tests to assess the significance of any association.

The R package ‘CCA’ was used for canonical correlation analysis (CCA) to find linear combinations of chemokines and cytokines with maximal correlation to a linear combination of the up-regulated miRNAs associated with cell death. To prevent overfitting, we only included cytokines and chemokines with individual correlation to any of these miRNAs > 0.4 in absolute value. We developed a model using the 43 samples from 12 patients (chosen randomly) with severe disease, using Wilks’ Lambda with Rao’s F-approximation to assess the significance of correlations. We used the data for the remaining 5 severe patients (17 observations) as an independent test set.

For all correlations between individual variables, Spearman’s rank-order correlation was used. Where there were missing values for clinical data, correlations were calculated over all patients for which values were available. Heatmaps showing multiple correlations were created using the package ‘pheatmap’ in R with thresholds for the minimum absolute values shown chosen to limit the size of the heatmaps.

## Data Availability

All data are available.

## AUTHOR CONTRIBUTIONS

JW, DL, NS, and PMK designed and supervised the study and performed data analysis. SJ and DK performed miRNA and cytokine/chemokine profiling experiments, respectively. KN performed miRNA profiling bioinformatics analyses. ERM, MM, SBK, JEK, CJ, KF, JRG and TH designed the sample collection study and performed clinical tissue processing, data collection and analysis. SK and MR performed clinical data analysis. AS performed sample collection and biobanking and contributed to data interpretation, presentation of clinical data and paper construction. CIRCO contributed to study design, patient consent and sample collection, and manuscript preparation. CIRCO performed tissue processing, data collection, and data analysis. DL, NS, and JW co-wrote the manuscript.

## ACKNOWLEDGEMENTS

The study was funded by the UKRI MRC/NIHR award to the UK Corovanirus Immunology Consortium (UK-CIC, MR/V028448/1). Sample collection was supported by the NIHR Manchester Biomedical Research Centre (TH and AS) and 3M Global Giving award (JRG and TH). JRG is supported by a senior fellowship by The Kennedy Trust for Rheumatology Research. This report is independent research supported by the North West Lung Centre Charity and the NIHR Manchester Clinical Research Facility at Wythenshawe Hospital. We acknowledge the Manchester Allergy, Respiratory, and Thoracic Surgery Biobank and thank the study participants for their contribution. The views expressed in this publication are those of the authors and not necessarily those of the NHS, the National Institute for Health Research, or the Department of Health. See Supplemental Acknowledgments for CIRCO consortium details. We thank staff at the Imaging and Cytometry Lab in the University of York Bioscience Technology Facility for technical support and advice.

