## Supplemental Figures for "Integrated miRNA/cytokine/chemokine profiling reveals immunopathological step changes associated with COVID-19 severity"

**Supplemental Figure 1:** miRNA signatures of COVID-19

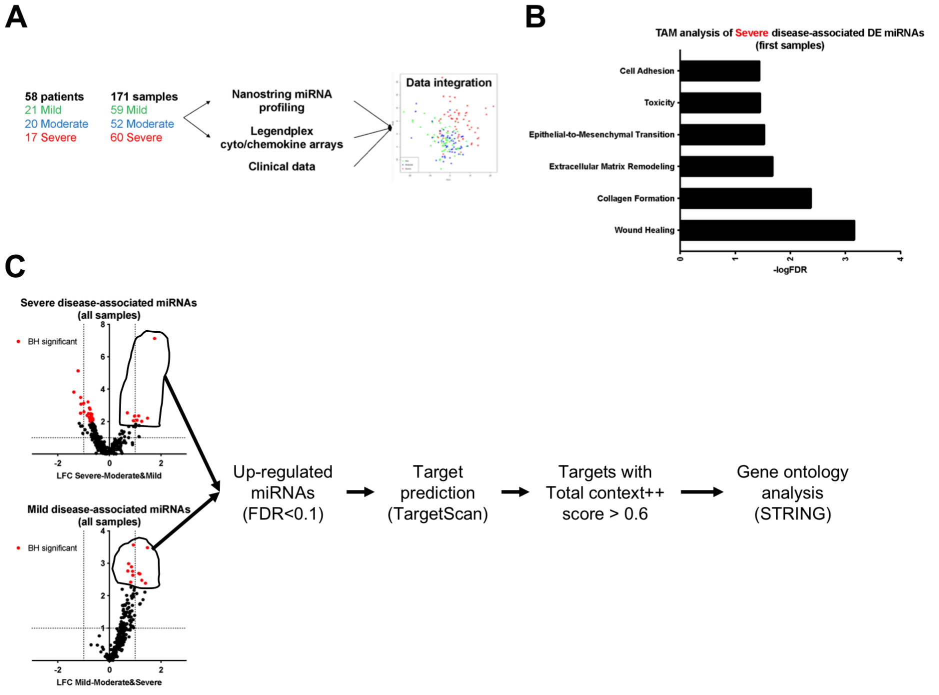


**Supplemental Figure 1: Study design and miRNA profiling of hospitalised patients with COVID-19**

**A.** Study schematic overview.

**B.** Significantly over-represented functional terms within DE miRNAs in severe cases, including only first samples (corresponding to Figure 1C).

**C.** Schematic outlining selection of top predicted targets of miRNAs up-regulated in severe or mild COVID-19 samples. TargetScan was used to identify all targets for all selected miRNAs (circled in the volcano plots) and high confidence targets (Total context++ score > 0.6) were selected for gene ontology analysis.

**Supplemental Figure 2:** Cytokine and chemokine signatures of COVID-19

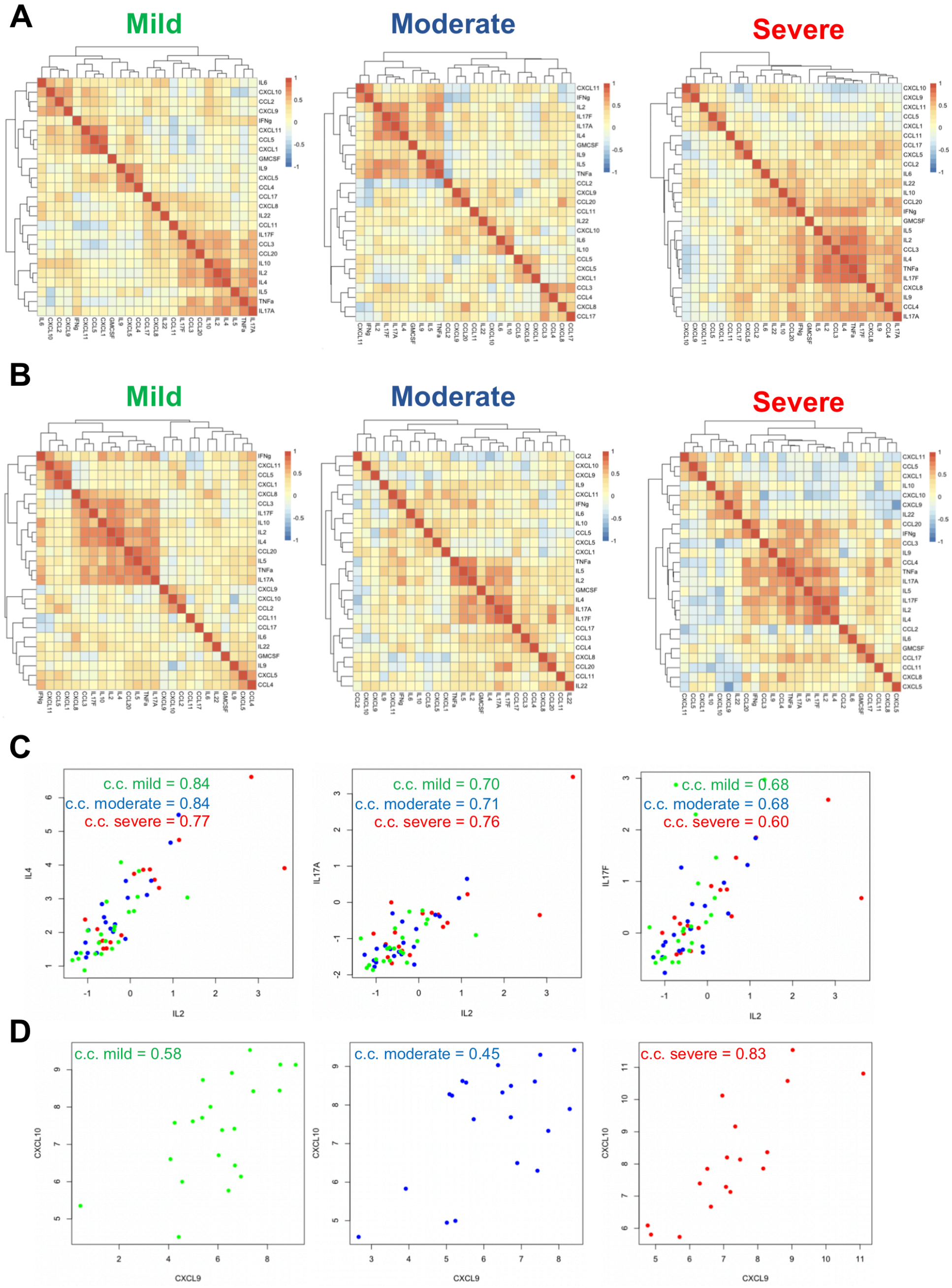


**Supplemental Figure 2: Cytokine and chemokine signatures of COVID-19**

**A.** Heatmaps showing Spearman correlation coefficients (c.c.) between cytokines and chemokines for mild, moderate, and severe groups, using all samples individually.

**B.** As in A, but only using measurements from first available samples for each patient.

**C.** Plots of IL2 against IL4, IL17A, and IL17F levels in mild (green), moderate (blue), and severe (red) patients (using. Correlation coefficients shown at the top of each dot plot.

**D.** Plots of CXCL9 against CXCL10 levels in mild (green), moderate (blue), and severe (red) patients. Correlation coefficients (c.c.) also shown.

**Supplemental Figure 3:** Integration of miRNA, cytokine, and chemokine signatures

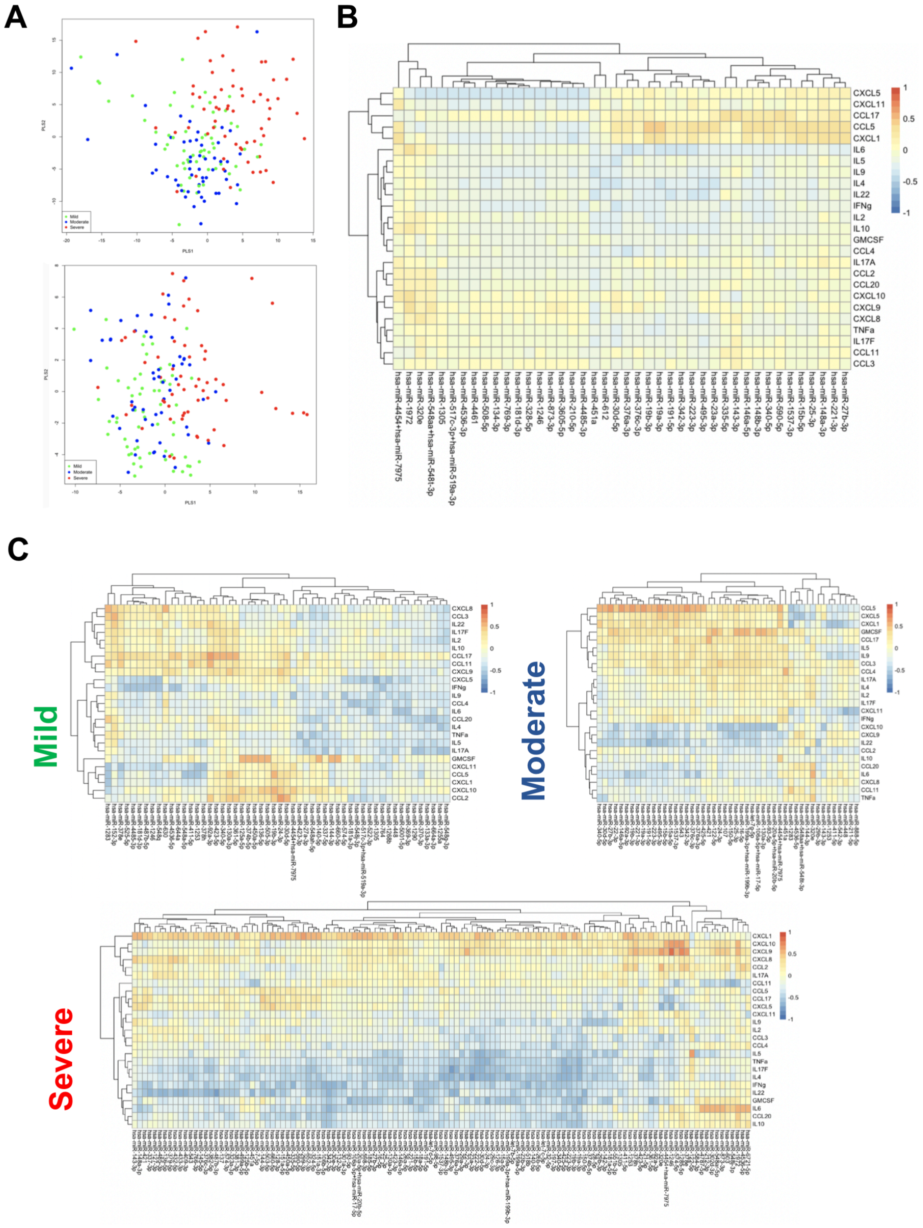


**Supplemental Figure 3: Integration of miRNA, cytokine, and chemokine signatures**

**A.** Scores plots for the first two latent variables from partial least squares regression (PLSR) obtained using DE miRNA measurements (left) or cytokine and chemokine values (right), coloured by severity group (mild, moderate and severe).

**B.** Heatmaps showing Spearman correlation coefficients (c.c.) between miRNAs and cytokines and chemokines. The miRNAs shown have a correlation greater than 0.3 in absolute value with at least one cytokine or chemokine. No values reach absolute c.c. of 0.4. Correlations are calculated over all severity groups after averaging values for any timepoints available for each patient.

**C.** Heatmaps showing Spearman correlation coefficients (c.c.) between miRNAs and cytokines and chemokines for mild, moderate, and severe groups. For each patient, data for all available timepoints are averaged. All miRNAs with c.c. > 0.5 in absolute value with any cytokine or chemokine are included.

### **Supplemental Figure 4:** Top correlated miRNAs with clinical parameters

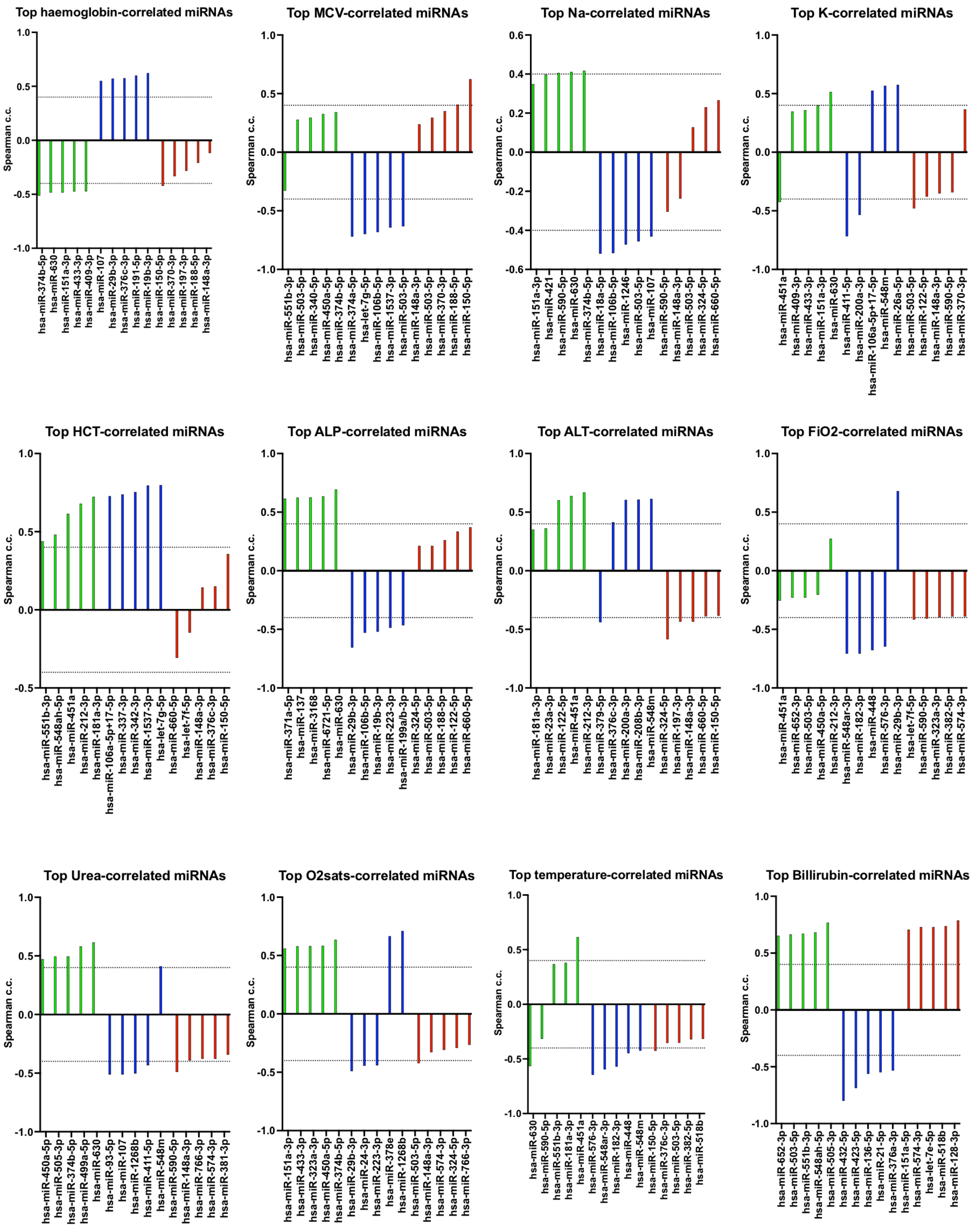


**Supplemental Figure 4: Top correlated miRNAs with clinical parameters**

Spearman correlation coefficients for top 5 correlated miRNAs with indicated clinical parameters in mild (green), moderate (blue), and severe (red) groups.

### SUPPLEMENTAL TABLES

**Supplemental Table 1: Study cohort characteristics and data: Age, sex, and ethnicity data for the whole cohort and per severity group.**

**Supplemental Table 2: Study cohort characteristics and data: Clinical measurements for the whole cohort and per severity group.**

**Supplemental Table 3: Study cohort characteristics and data: Cohort comorbidities and symptoms.**

**Supplemental Table 4: Study cohort characteristics and data: Sampling information for the whole cohort and per severity group.**

**Supplemental Table 5: MiRNA profiling of COVID-19 patients: Severe vs Mild and Moderate.**

Statistics for all 280 detected miRNAs for comparisons (Severe vs Mild and Moderate) including all samples (corresponding to Figure 1A) and first available (corresponding to Figure 1C) samples for each patient.

**Supplemental Table 6: MiRNA profiling of COVID-19 patients: Severe vs Mild and Moderate.**

Statistics for all 280 detected miRNAs for comparisons (Mild vs Moderate and Severe) including all samples (corresponding to Figure 2A) and first available (corresponding to Figure 2C) samples for each patient.

**Supplemental Table 7: Cytokine and chemokine profiling of COVID-19 patients**

Statistics for comparisons including all samples or only first available sample for each patient. Mann Whitney P values before and after Benjamini-Hochberg correction shown.

**Supplemental Table 8: Strongest miRNA correlations for severity associated cytokines and chemokines**

The 5 miRNAs with the strongest Spearman correlation to each cytokine/chemokine found to be significantly different in Mann-Whitney tests between either the severe and mild/moderate groups or the mild and moderate/severe groups. Negative correlations shown in red. Coloured cells indicate miRNAs that are within the top 5 correlations for more than one cytokine/chemokine. Data shown for mild, moderate, or severe cases.

**Supplemental Table 9: Strongest cytokine and chemokine correlations for Cell Death-associated miRNAs**

The 5 cytokines/chemokines with the strongest Spearman correlation (over severe samples) to each miRNA associated with Cell Death (see **Figure 1E**). Negative correlations are highlighted in red text.

**SUPPLEMENTAL ACKNOLWEDGEMENTS**

The CIRCO cohort members: Rohan Ahmed, Miriam Avery, Katharine Birchall, Evelyn Charsley, Alistair Chenery, Christine Chew, Richard Clark, Emma Connolly, Karen Connolly, Simon Dawson, Laura Durrans, Hannah Durrington, Jasmine Egan, Timothy Felton, Claire Fox, Helen Francis, Miriam Franklin, Susannah Glasgow, Nicola Godfrey, Kathryn J Gray, Seamus Grundy, Jacinta Guerin, Pamela Hackney, Chantelle Hayes, Emma Hardy, Jade Harris, Anu John, Bethany Jolly, Verena Kästele, Gina Kerry, Sara Kirkham, Sylvia Lui, Lijing Lin, Alex G Mathioudakis, Joanne Mitchell, Clare Moizer, Katrina Moore, Stuart Moss, Syed Murtuza Baker, Rob Oliver, Grace Padden, Christina Parkinson, Michael Phuycharoen, Magnus Rattray, Ananya Saha, Barbora Salcman, Nicholas A Scott, Seema Sharma, Jane Shaw, Joanne Shaw, Elizabeth Shepley, Lara Smith, Simon Stephan, Ruth Stephens, Gael Tavernier, Rhys Tudge, Louis Wareing, Roanna Warren, Thomas Williams, Lisa Willmore, Mehwish Younas.
