## Supplemental Tables for "Integrated miRNA/cytokine/chemokine profiling reveals immunopathological step changes associated with COVID-19 severity"

**Supplemetary Table 1: Study cohort characteristics and data; Age, Sex, Ethnicity.**
**Supplemetary Table 2: Study cohort characteristics and data; Clinical measurements.**
**Supplemetary Table 3: Study cohort characteristics and data; Comorbidities and symptoms.**
**Supplemetary Table 4: Study cohort characteristics and data; Sampling information.**
**Supplementary Table 5: MiRNA profiling of COVID-19 patients: Severe vs Mild and Moderate.**
**Supplementary Table 6: MiRNA profiling of COVID-19 patients: Mild vs Moderate and Severe.**
**Supplementary Table 7: Cytokine and chemokine profiling of COVID-19 patients.**
**Supplementary Table 8: Strongest miRNA correlations for severity associated cytokines and chemokines.**
**Supplementary Table 9: Strongest cytokine and chemokine correlations for Cell Death-associated miRNAs.**

| **Age** | All | Mild | Moderate | Severe |
|---|---|---|---|---|
| Median | 63 | 59 | 62 | 64 |
| Minimum | 22 | 22 | 27 | 31 |
| Maximum | 97 | 84 | 90 | 97 |

| | **Sex (F/M)** | **Ethnicity (BAME)** |
|---|---|---|
| All | 22/36 | 6 |
| Mild | 9/12 | 3 |
| Moderate | 6/14 | 0 |
| Severe | 7/10 | 3 |

| | BMI | admission Fi02 | admission o2 sats | admission RR | admission temperature | admission HR | admission BP systolic | admission BP diastolic | White blood cell count (×10^9/liter) | Lymphocytes (×10^9/liter) | Neutrophils (×10^9/liter) | Eosinophils (×10^9/liter) | Monocytes (×10^9/liter) | Basophils (×10^9/liter) |
|---|---|---|---|---|---|---|---|---|---|---|---|---|---|---|
| Median (all) | 28.3 | 21 | 96 | 19.5 | 37.1 | 91.5 | 126 | 74 | 7.25 | 1.1 | 5.4 | 0 | 0.4 | 0 |
| Minimum (all) | 13.6 | 21 | 28 | 15 | 35.9 | 60 | 97 | 33 | 3.7 | 0.2 | 0.5 | 0 | 0 | 0 |
| Maximum (all) | 42 | 100 | 100 | 38 | 39.5 | 126 | 194 | 113 | 27.1 | 3.1 | 22.8 | 0.5 | 1.6 | 0.1 |
| Median (mild) | 27.8 | 21 | 97 | 19 | 37 | 86 | 126 | 68 | 6.9 | 1.1 | 4.3 | 0 | 0.3 | 0 |
| Minimum (mild) | 20.3 | 21 | 92 | 16 | 35.9 | 60 | 99 | 33 | 4.1 | 0.2 | 0.5 | 0 | 0 | 0 |
| Maximum (mild) | 37.3 | 35 | 100 | 28 | 39.4 | 126 | 160 | 95 | 27.1 | 3.1 | 22.8 | 0.5 | 0.8 | 0.1 |
| Median (moderate) | 28.7 | 24 | 95 | 18.5 | 37.4 | 87.5 | 123 | 72.5 | 7.35 | 1.3 | 5.35 | 0 | 0.5 | 0 |
| Minimum (moderate) | 13.6 | 21 | 28 | 16 | 36.1 | 63 | 97 | 35 | 4.2 | 0.6 | 2.8 | 0 | 0.2 | 0 |
| Maximum (moderate) | 42 | 60 | 100 | 28 | 39.5 | 120 | 156 | 95 | 19 | 2.6 | 17 | 0.2 | 1.6 | 0.1 |
| Median (severe) | 27.7 | 28 | 95 | 22 | 37.7 | 99 | 133 | 82 | 8.2 | 0.9 | 6.2 | 0 | 0.3 | 0 |
| Minimum (severe) | 21.3 | 21 | 80 | 15 | 36.2 | 60 | 116 | 45 | 3.7 | 0.4 | 2.8 | 0 | 0.16 | 0 |
| Maximum (severe) | 33.1 | 100 | 99 | 38 | 39.1 | 123 | 194 | 113 | 16.2 | 1.72 | 14.7 | 0.28 | 1.1 | 0.1 |

| | Haemoglobin (g/litre) | MCV (femtoliters) | Haematocrit | Platelets (×109/liter) | Na (mEq/Litre) | K (mEq/Litre) | Urea (mg/dL) | Creatinine (micromol/litre)) | CRP (mg/liter) | Albumin (g/liter) | Globulin (g/litre) | Alkaline phosphatase (U/liter) | Alanine aminotransferase (U/liter) | Bilirubin (μM) |
|---|---|---|---|---|---|---|---|---|---|---|---|---|---|---|
| Median (all) | 132 | 87.9 | 0.395 | 242.5 | 136 | 4 | 6 | 80 | 71 | 39 | 28 | 76.5 | 41.5 | 10 |
| Minimum (all) | 82 | 61.2 | 0.232 | 78 | 128 | 2.8 | 2.2 | 41 | 3.3 | 17 | 22 | 48 | 14 | 3 |
| Maximum (all) | 173 | 107 | 0.5 | 625 | 145 | 5.1 | 25.3 | 468 | 356.5 | 51 | 40 | 241 | 181 | 298 |
| Median (mild) | 130 | 88.05 | 0.395 | 247 | 137 | 4 | 5.7 | 75.5 | 48.6 | 38 | 27.5 | 79 | 52 | 9.5 |
| Minimum (mild) | 86 | 78.8 | 0.29 | 153 | 133 | 3.3 | 2.2 | 41 | 3.3 | 25 | 22 | 52 | 18 | 4 |
| Maximum (mild) | 173 | 98.9 | 0.496 | 625 | 143 | 4.6 | 16.5 | 157 | 193 | 51 | 30 | 181 | 139 | 28 |
| Median (moderate) | 123.5 | 89 | 0.37 | 262 | 134 | 3.9 | 5.8 | 78.5 | 68 | 37.5 | 26 | 67.5 | 35 | 11 |
| Minimum (moderate) | 82 | 79.7 | 0.232 | 78 | 128 | 3.2 | 2.7 | 63 | 14.6 | 17 | 22 | 48 | 14 | 3 |
| Maximum (moderate) | 155 | 107 | 0.45 | 583 | 145 | 4.4 | 25.2 | 144 | 216 | 48 | 31 | 177 | 181 | 298 |
| Median (severe) | 143 | 84.75 | 0.43 | 173 | 134.5 | 4 | 8.55 | 90 | 131.5 | 39 | 30 | 85.5 | 36 | 10 |
| Minimum (severe) | 98 | 61.2 | 0.28 | 124 | 131 | 2.8 | 2.9 | 52 | 23 | 22 | 23 | 52 | 25 | 5 |
| Maximum (severe) | 168 | 93.4 | 0.5 | 447 | 139 | 5.1 | 25.3 | 468 | 356.5 | 47 | 40 | 241 | 181 | 26 |

| | Hypertension | Ischaemic Heart Diseae | Chronic lung disease (non asthma) | Asthma | Diabetes | Other comorbidities | Fever | Cough | Shortness of breath | Myalgia | Fatigue | GI_symptoms | Headache |
|---|---|---|---|---|---|---|---|---|---|---|---|---|---|
| Number (all) | 20 | 3 | 8 | 13 | 13 | 29 | 27 | 31 | 37 | 13 | 13 | 13 | 5 |
| Percentage (all) | 34.5 | 5.2 | 13.8 | 22.4 | 22.4 | 50.0 | 46.6 | 53.4 | 63.8 | 22.4 | 22.4 | 22.4 | 8.6 |
| Number (mild) | 4 | 2 | 2 | 5 | 5 | 10 | 10 | 10 | 9 | 4 | 3 | 5 | 3 |
| Percentage (mild) | 19.0 | 9.5 | 9.5 | 23.8 | 23.8 | 47.6 | 47.6 | 47.6 | 42.9 | 19.0 | 14.3 | 23.8 | 14.3 |
| Number (moderate) | 6 | 0 | 4 | 5 | 3 | 10 | 6 | 10 | 13 | 2 | 4 | 3 | 2 |
| Percentage (moderate) | 30 | 0 | 20 | 25 | 15 | 50 | 30 | 50 | 65 | 10 | 20 | 15 | 10 |
| Number (severe) | 10 | 1 | 2 | 3 | 5 | 9 | 11 | 11 | 15 | 7 | 6 | 5 | 0 |
| Percentage (severe) | 58.8 | 5.9 | 11.8 | 17.6 | 29.4 | 52.9 | 64.7 | 64.7 | 88.2 | 41.2 | 35.3 | 29.4 | 0.0 |

| | Illness onset to admission (days) | Illnes onset to last sample (days) | Day recruited | Number of samples per patient |
|---|---|---|---|---|
| Median (all) | 2 | 5 | 3 | 2 |
| Minimum (all) | 1 | 1 | 0 | 1 |
| Maximum (all) | 11 | 146 | 28 | 8 |
| Median (mild) | 1 | 4 | 2 | 2 |
| Minimum (mild) | 1 | 1 | 1 | 1 |
| Maximum (mild) | 9 | 146 | 24 | 8 |
| Median (moderate) | 2 | 4.5 | 3 | 2 |
| Minimum (moderate) | 1 | 1 | 1 | 1 |
| Maximum (moderate) | 4 | 10 | 28 | 6 |
| Median (severe) | 1 | 8 | 3 | 3 |
| Minimum (severe) | 1 | 1 | 0 | 1 |
| Maximum (severe) | 11 | 112 | 13 | 8 |

| Number of patients with all timepoints within 2 weeks of illness onset |
|---|
| 48 (82.8%) |
| 18 (85.7%) |
| 20 (100%) |
| 10 (58.8%) |

**All samples**

| miRNA | BH adjusted MW test P value - | LFC Severe- |
|---|---|---|
| hsa-miR-4454+hs | 2.08E-05 | 1.754 |
| hsa-miR-451a | 0.001031842 | -1.218 |
| hsa-miR-323a-3p | 0.013965413 | -1.379 |
| hsa-miR-188-5p | 0.023057565 | -1.113 |
| hsa-miR-432-5p | 0.033685431 | -1.105 |
| hsa-miR-433-3p | 0.033685431 | -0.994 |
| hsa-miR-371a-5p | 0.033685431 | -0.831 |
| hsa-miR-3144-3p | 0.050693735 | -0.771 |
| hsa-miR-302d-3p | 0.054122596 | -0.753 |
| hsa-miR-134-3p | 0.064448438 | -0.719 |
| hsa-miR-612 | 0.064448438 | -1.118 |
| hsa-let-7a-5p | 0.064448438 | 0.697 |
| hsa-miR-548ah-5 | 0.064448438 | -0.988 |
| hsa-miR-4787-5p | 0.064448438 | -0.808 |
| hsa-miR-764 | 0.064448438 | -0.684 |
| hsa-miR-4536-5p | 0.06574454 | -0.867 |
| hsa-miR-627-3p | 0.06574454 | -0.689 |
| hsa-miR-212-3p | 0.06574454 | -0.721 |
| hsa-miR-4516 | 0.06574454 | 0.977 |
| hsa-miR-630 | 0.06574454 | 1.134 |
| hsa-miR-651-5p | 0.068550094 | -0.730 |
| hsa-miR-4536-3p | 0.070172271 | -0.779 |
| hsa-miR-29b-3p | 0.074844062 | 1.481 |
| hsa-miR-181d-3p | 0.076825352 | -0.648 |
| hsa-miR-922 | 0.080506898 | -0.749 |
| hsa-miR-1278 | 0.081671398 | -0.675 |
| hsa-miR-328-5p | 0.081671398 | -0.640 |
| hsa-miR-181a-3p | 0.081671398 | -0.652 |
| hsa-let-7f-5p | 0.081671398 | 1.070 |
| hsa-miR-574-5p | 0.081671398 | 0.930 |
| hsa-miR-508-5p | 0.083655497 | -0.753 |
| hsa-miR-24-3p | 0.083759305 | 1.254 |
| hsa-miR-210-5p | 0.097391027 | -0.686 |
| hsa-miR-519c-3p | 0.100803588 | -0.663 |
| hsa-miR-495-3p | 0.102965176 | -1.168 |
| hsa-miR-203a-5p | 0.11003313 | -0.661 |
| hsa-miR-29a-3p | 0.11003313 | 1.022 |
| hsa-miR-409-3p | 0.111539467 | -0.607 |
| hsa-miR-382-5p | 0.111716123 | -1.110 |
| hsa-miR-381-3p | 0.111716123 | -0.595 |
| hsa-miR-942-5p | 0.111716123 | -0.605 |
| hsa-miR-7-5p | 0.111716123 | -0.669 |
| hsa-miR-144-3p | 0.111716123 | -1.056 |
| hsa-miR-548m | 0.111716123 | -0.677 |
| hsa-miR-873-3p | 0.111716123 | -0.769 |
| hsa-miR-22-3p | 0.111716123 | 0.937 |
| hsa-miR-26a-5p | 0.111716123 | 1.011 |
| hsa-miR-1249-3p | 0.111716123 | -0.556 |
| hsa-miR-3605-5p | 0.111716123 | -0.699 |
| hsa-miR-548ar-5 | 0.111716123 | -0.629 |
| hsa-miR-1283 | 0.111716123 | 1.147 |
| hsa-miR-320e | 0.111716123 | 0.748 |
| hsa-miR-6721-5p | 0.12090669 | -0.721 |
| hsa-miR-548j-3p | 0.134056413 | -0.556 |
| hsa-miR-5001-5p | 0.140658662 | -0.628 |
| hsa-miR-4461 | 0.143367858 | -0.582 |
| hsa-miR-376a-3p | 0.157893359 | -0.945 |
| hsa-miR-152-3p | 0.168506337 | -0.524 |
| hsa-miR-301b-3p | 0.168506337 | -0.662 |
| hsa-miR-4284 | 0.168506337 | -0.576 |
| hsa-miR-148a-3p | 0.173816861 | 0.478 |
| hsa-miR-499a-5p | 0.175801268 | -0.613 |
| hsa-miR-3613-3p | 0.18205133 | -0.594 |
| hsa-miR-127-3p | 0.187768192 | -0.585 |
| hsa-miR-1285-5p | 0.187768192 | 0.569 |
| hsa-miR-526b-5p | 0.189253777 | -0.542 |
| hsa-miR-937-3p | 0.189321245 | -0.555 |
| hsa-miR-543 | 0.200092597 | -0.854 |
| hsa-miR-210-3p | 0.200092597 | -0.578 |
| hsa-miR-221-3p | 0.200092597 | 0.771 |
| hsa-miR-376c-3p | 0.211536036 | -1.032 |
| hsa-miR-4707-5p | 0.211536036 | -0.492 |
| hsa-miR-27b-3p | 0.211536036 | 0.518 |
| hsa-let-7g-5p | 0.227194758 | 0.405 |
| hsa-miR-208b-3p | 0.227476289 | -0.698 |
| hsa-miR-208a-3p | 0.250446549 | -0.572 |
| hsa-miR-525-5p | 0.250446549 | -0.597 |
| hsa-miR-487b-5p | 0.250446549 | -0.558 |
| hsa-miR-615-3p | 0.250446549 | -0.706 |
| hsa-miR-10b-5p | 0.282185575 | -0.688 |
| hsa-miR-650 | 0.282185575 | -0.436 |
| hsa-miR-107 | 0.287331845 | 0.592 |
| hsa-miR-1224-3p | 0.287331845 | -0.327 |
| hsa-miR-576-3p | 0.287331845 | -0.516 |
| hsa-miR-603 | 0.287331845 | -0.451 |
| hsa-miR-1228-3p | 0.287331845 | -0.478 |
| hsa-miR-1972 | 0.287331845 | 1.148 |
| hsa-miR-769-5p | 0.297558661 | -0.453 |
| hsa-miR-1268b | 0.297558661 | -0.635 |
| hsa-miR-142-3p | 0.301483446 | 0.746 |
| hsa-miR-769-3p | 0.301483446 | -0.429 |
| hsa-miR-18b-5p | 0.301483446 | -0.319 |
| hsa-miR-136-5p | 0.301483446 | -0.393 |
| hsa-miR-301a-5p | 0.314329254 | -0.652 |
| hsa-miR-1253 | 0.314329254 | 0.456 |
| hsa-miR-485-3p | 0.317251023 | -0.434 |
| hsa-miR-125b-5p | 0.322306663 | -0.370 |
| hsa-miR-598-3p | 0.329562257 | 0.435 |
| hsa-miR-377-3p | 0.332624776 | -0.406 |
| hsa-miR-20a-5p+l | 0.338969224 | 0.210 |
| hsa-miR-19b-3p | 0.345229787 | 0.228 |
| hsa-miR-548ar-3 | 0.345229787 | -0.475 |
| hsa-miR-626 | 0.345229787 | -0.549 |
| hsa-miR-130a-3p | 0.355123355 | 0.373 |
| hsa-miR-1973 | 0.358399074 | -0.509 |
| hsa-miR-1295a | 0.358399074 | -0.665 |
| hsa-miR-340-5p | 0.375446935 | -0.427 |
| hsa-miR-10a-5p | 0.375446935 | -0.402 |
| hsa-miR-423-3p | 0.393067491 | 0.577 |
| hsa-miR-106a-5p | 0.395552687 | 0.222 |
| hsa-miR-93-5p | 0.395552687 | 0.307 |
| hsa-miR-412-3p | 0.395552687 | -0.412 |
| hsa-miR-888-5p | 0.395552687 | 0.413 |
| hsa-miR-148b-3p | 0.401815626 | -0.438 |
| hsa-miR-106b-5p | 0.401815626 | 0.583 |
| hsa-miR-362-5p | 0.401815626 | -0.478 |
| hsa-miR-369-3p | 0.415250697 | -0.485 |
| hsa-miR-548v | 0.421670601 | -0.307 |
| hsa-miR-548aa+h | 0.421670601 | -0.196 |
| hsa-miR-126-3p | 0.42183702 | 0.312 |
| hsa-miR-19a-3p | 0.434618965 | 0.188 |
| hsa-miR-370-3p | 0.434618965 | -0.576 |
| hsa-miR-23b-3p | 0.446184522 | 0.535 |
| hsa-let-7d-5p | 0.455357052 | 0.895 |
| hsa-miR-29c-3p | 0.46334204 | 0.633 |
| hsa-miR-421 | 0.478036109 | 0.386 |
| hsa-miR-939-5p | 0.487602915 | -0.442 |
| hsa-miR-361-5p | 0.494574036 | 0.428 |
| hsa-miR-454-3p | 0.501600538 | -0.428 |
| hsa-miR-4485-3p | 0.509448963 | -0.294 |
| hsa-miR-301b-5p | 0.509448963 | -0.505 |
| hsa-miR-597-5p | 0.509448963 | -0.405 |
| hsa-miR-1185-5p | 0.509448963 | -0.503 |
| hsa-miR-182-3p | 0.510379719 | -0.415 |
| hsa-miR-137 | 0.510379719 | -0.412 |
| hsa-miR-378e | 0.510379719 | 0.107 |
| hsa-miR-644a | 0.510726817 | -0.375 |
| hsa-miR-505-3p | 0.518287886 | -0.218 |
| hsa-miR-660-5p | 0.518287886 | 0.259 |
| hsa-miR-132-3p | 0.521416119 | 0.294 |
| hsa-miR-374a-5p | 0.52181426 | 0.898 |
| hsa-miR-125a-5p | 0.522231233 | 0.898 |
| hsa-miR-191-5p | 0.541987095 | 0.450 |
| hsa-miR-363-3p | 0.541987095 | 0.169 |
| hsa-miR-378d | 0.541987095 | -0.531 |
| hsa-miR-517c-3p | 0.541987095 | -0.312 |
| hsa-miR-3147 | 0.541987095 | -0.385 |
| hsa-miR-1290 | 0.541987095 | -0.478 |
| hsa-miR-1197 | 0.541987095 | -0.315 |
| hsa-miR-301a-3p | 0.545261472 | 0.121 |
| hsa-miR-585-3p | 0.554117734 | -0.482 |
| hsa-miR-200a-3p | 0.558832694 | -0.462 |
| hsa-miR-584-3p | 0.560763579 | -0.458 |
| hsa-miR-185-5p | 0.596988512 | 0.105 |
| hsa-miR-425-5p | 0.597487707 | 0.275 |
| hsa-miR-15a-5p | 0.600766161 | 0.339 |
| hsa-miR-15b-5p | 0.600766161 | 0.424 |
| hsa-miR-514b-5p | 0.600766161 | -0.407 |
| hsa-miR-450a-5p | 0.600766161 | -0.186 |
| hsa-miR-767-5p | 0.600766161 | -0.484 |
| hsa-miR-498 | 0.600766161 | 0.173 |
| hsa-miR-582-5p | 0.6033964 | -0.381 |
| hsa-miR-4488 | 0.6033964 | -0.349 |
| hsa-miR-1302 | 0.606883058 | -0.480 |
| hsa-miR-16-5p | 0.616777149 | 0.147 |
| hsa-miR-18a-5p | 0.616777149 | 0.428 |
| hsa-miR-5010-3p | 0.65117882 | -0.250 |
| hsa-miR-411-5p | 0.654746174 | 0.168 |
| hsa-miR-652-3p | 0.65533668 | -0.088 |
| hsa-miR-379-5p | 0.673017626 | 0.100 |
| hsa-miR-197-5p | 0.673017626 | -0.446 |
| hsa-miR-532-3p | 0.676622656 | -0.456 |
| hsa-miR-4647 | 0.693913536 | -0.260 |
| hsa-miR-514a-3p | 0.694491521 | -0.296 |
| hsa-miR-199a-5p | 0.695681316 | 0.114 |
| hsa-miR-891a-5p | 0.695681316 | -0.418 |
| hsa-miR-186-5p | 0.70208037 | -0.465 |
| hsa-miR-27a-3p | 0.70208037 | 0.144 |
| hsa-miR-211-5p | 0.70208037 | -0.422 |
| hsa-miR-1255a | 0.724078959 | -0.335 |
| hsa-let-7b-5p | 0.725903735 | -0.359 |
| hsa-miR-548a-5p | 0.725903735 | -0.371 |
| hsa-miR-34a-5p | 0.725903735 | -0.372 |
| hsa-miR-145-5p | 0.729587501 | 0.044 |
| hsa-miR-187-3p | 0.738497667 | -0.356 |
| hsa-miR-222-3p | 0.738497667 | 0.329 |
| hsa-miR-378f | 0.741297791 | -0.326 |
| hsa-miR-551b-3p | 0.741297791 | -0.186 |
| hsa-miR-593-3p | 0.752657556 | -0.160 |
| hsa-miR-30e-3p | 0.754576829 | 0.128 |
| hsa-miR-28-5p | 0.754576829 | 0.357 |
| hsa-miR-933 | 0.754576829 | -0.268 |
| hsa-miR-146a-5p | 0.755893505 | 0.235 |
| hsa-miR-33b-5p | 0.755893505 | -0.291 |
| hsa-miR-92a-3p | 0.757729041 | 0.284 |
| hsa-miR-518b | 0.757729041 | 0.417 |
| hsa-miR-98-5p | 0.757729041 | 0.429 |
| hsa-miR-503-5p | 0.757729041 | -0.243 |
| hsa-miR-487a-3p | 0.757729041 | -0.211 |
| hsa-miR-542-3p | 0.757729041 | -0.401 |
| hsa-miR-513a-3p | 0.757729041 | 0.066 |
| hsa-miR-4286 | 0.777163629 | 0.072 |
| hsa-miR-574-3p | 0.777163629 | -0.011 |
| hsa-miR-302a-3p | 0.782377957 | -0.216 |
| hsa-miR-625-5p | 0.783070267 | 0.133 |
| hsa-miR-512-5p | 0.788279847 | -0.287 |
| hsa-miR-25-5p | 0.796502555 | -0.248 |
| hsa-miR-423-5p | 0.79869118 | -0.137 |
| hsa-miR-548e-5p | 0.802385675 | -0.295 |
| hsa-miR-365a-3p | 0.803068532 | -0.438 |
| hsa-miR-1246 | 0.812776931 | -0.251 |
| hsa-let-7c-5p | 0.81975346 | -0.262 |
| hsa-miR-183-5p | 0.81975346 | -0.329 |
| hsa-miR-30e-5p | 0.81975346 | -0.015 |
| hsa-miR-522-3p | 0.81975346 | -0.313 |
| hsa-miR-497-5p | 0.81975346 | 0.108 |
| hsa-miR-30b-5p | 0.81975346 | 0.147 |
| hsa-miR-448 | 0.81975346 | 0.039 |
| hsa-miR-563 | 0.81975346 | -0.162 |
| hsa-miR-1537-3p | 0.823412561 | -0.002 |
| hsa-miR-181a-5p | 0.856389915 | 0.135 |
| hsa-miR-337-5p | 0.856389915 | -0.015 |
| hsa-miR-130b-3p | 0.856389915 | -0.013 |
| hsa-miR-3168 | 0.856389915 | -0.200 |
| hsa-miR-1185-2-3 | 0.856389915 | 0.000 |
| hsa-miR-1262 | 0.857032958 | -0.297 |
| hsa-miR-642a-5p | 0.858327205 | -0.216 |
| hsa-miR-140-5p | 0.858327205 | 0.463 |
| hsa-miR-151a-3p | 0.865494507 | -0.012 |
| hsa-miR-766-3p | 0.865494507 | 0.079 |
| hsa-miR-155-5p | 0.873465507 | 0.158 |
| hsa-miR-510-3p | 0.875554592 | -0.193 |
| hsa-miR-324-3p | 0.886426839 | -0.230 |
| hsa-miR-122-5p | 0.889681045 | 0.108 |
| hsa-miR-28-3p | 0.889681045 | -0.096 |
| hsa-miR-1296-3p | 0.889681045 | -0.228 |
| hsa-miR-23a-3p | 0.890279502 | -0.308 |
| hsa-miR-151a-5p | 0.89968296 | -0.095 |
| hsa-miR-26b-5p | 0.899882323 | 0.005 |
| hsa-miR-378h | 0.899882323 | -0.176 |
| hsa-miR-299-5p | 0.899882323 | -0.008 |
| hsa-miR-95-3p | 0.910535546 | -0.052 |
| hsa-miR-487b-3p | 0.918284025 | -0.088 |
| hsa-miR-639 | 0.921695344 | -0.106 |
| hsa-miR-299-3p | 0.9241892 | -0.032 |
| hsa-miR-337-3p | 0.9241892 | -0.229 |
| hsa-miR-502-5p | 0.928994262 | 0.011 |
| hsa-miR-199a-3p | 0.941871905 | -0.159 |
| hsa-miR-25-3p | 0.941871905 | -0.027 |
| hsa-miR-802 | 0.941871905 | -0.227 |
| hsa-miR-590-5p | 0.945264564 | 0.067 |
| hsa-miR-570-3p | 0.945264564 | 0.009 |
| hsa-miR-3614-5p | 0.945264564 | -0.232 |
| hsa-miR-1305 | 0.945264564 | -0.024 |
| hsa-miR-627-5p | 0.945264564 | -0.052 |
| hsa-miR-495-5p | 0.945264564 | -0.082 |
| hsa-miR-199b-5p | 0.945264564 | -0.069 |
| hsa-miR-128-3p | 0.947610696 | 0.062 |
| hsa-miR-378i | 0.947610696 | -0.032 |
| hsa-miR-378g | 0.948089166 | -0.207 |
| hsa-miR-223-3p | 0.95130874 | -0.025 |
| hsa-miR-664a-3p | 0.95177728 | -0.145 |
| hsa-miR-30d-5p | 0.952711563 | -0.512 |
| hsa-miR-133a-3p | 0.952711563 | -0.148 |
| hsa-miR-150-5p | 0.958142743 | -0.048 |
| hsa-miR-548g-3p | 0.958142743 | -0.256 |
| hsa-miR-197-3p | 0.958142743 | 0.091 |
| hsa-miR-374b-5p | 0.9617154 | 0.492 |
| [illegible] | [illegible] | [illegible] |

**First samples only**

| miRNA | LFC Severe- | MW test P value - |
|---|---|---|
| hsa-miR-4454+hsa-miR-7975 | 1.831 | 0.000782642 |
| hsa-miR-1285-5p | 1.338 | 0.001633306 |
| hsa-miR-451a | -1.207 | 0.005085737 |
| hsa-miR-320e | 1.314 | 0.009652952 |
| hsa-miR-144-3p | -1.942 | 0.010491864 |
| hsa-miR-29b-3p | 1.840 | 0.010491864 |
| hsa-miR-1246 | 0.529 | 0.041264861 |
| hsa-miR-188-5p | -1.085 | 0.052967733 |
| hsa-miR-1972 | 1.584 | 0.059765487 |
| hsa-miR-151a-5p | -1.119 | 0.067255073 |
| hsa-miR-1253 | 0.875 | 0.075635026 |
| hsa-miR-574-5p | 0.947 | 0.081401797 |
| hsa-miR-4516 | 0.884 | 0.090964721 |
| hsa-let-7a-5p | 0.663 | 0.104621368 |
| hsa-miR-1305 | 0.825 | 0.108844013 |
| hsa-miR-764 | -0.567 | 0.116042746 |
| hsa-miR-888-5p | 0.912 | 0.116042746 |
| hsa-miR-627-5p | 0.860 | 0.139501997 |
| hsa-miR-132-3p | 0.890 | 0.142960867 |
| hsa-miR-212-3p | -0.525 | 0.15130377 |
| hsa-miR-450a-2-3p | 0.587 | 0.15262895 |
| hsa-miR-184 | 0.716 | 0.162776149 |
| hsa-miR-432-5p | -0.639 | 0.166452299 |
| hsa-miR-505-3p | -0.630 | 0.168032827 |
| hsa-miR-1228-3p | -0.513 | 0.171747284 |
| hsa-miR-939-5p | -0.430 | 0.177160671 |
| hsa-miR-502-5p | 0.899 | 0.177167334 |
| hsa-miR-183-5p | 0.557 | 0.177167334 |
| hsa-miR-24-3p | 1.264 | 0.184550602 |
| hsa-miR-323a-3p | -0.925 | 0.190309804 |
| hsa-miR-26a-5p | 0.967 | 0.196197546 |
| hsa-miR-4286 | 0.740 | 0.202214893 |
| hsa-miR-3147 | -0.537 | 0.206188709 |
| hsa-miR-495-3p | -0.978 | 0.208362859 |
| hsa-miR-802 | 0.680 | 0.208362859 |
| hsa-miR-548ah-5p | -0.629 | 0.208362859 |
| hsa-miR-340-5p | -0.679 | 0.212385585 |
| hsa-miR-6721-5p | -0.374 | 0.218715885 |
| hsa-miR-22-3p | 0.806 | 0.22105444 |
| hsa-miR-1255a | 0.620 | 0.227599807 |
| hsa-miR-152-3p | -0.457 | 0.231780636 |
| hsa-miR-590-5p | -0.808 | 0.234279297 |
| hsa-miR-548e-5p | 0.645 | 0.238516883 |
| hsa-miR-378e | 0.430 | 0.245382877 |
| hsa-miR-362-5p | -0.373 | 0.245390137 |
| hsa-miR-4461 | -0.383 | 0.259550566 |
| hsa-miR-25-5p | 0.468 | 0.266831664 |
| hsa-miR-142-3p | 0.602 | 0.269711831 |
| hsa-miR-130a-3p | 0.474 | 0.269711831 |
| hsa-miR-532-3p | 0.574 | 0.289543547 |
| hsa-miR-644a | -0.364 | 0.292617374 |
| hsa-miR-151a-3p | -0.672 | 0.300528841 |
| hsa-let-7g-5p | 0.373 | 0.300528841 |
| hsa-miR-1973 | -0.436 | 0.308578688 |
| hsa-miR-433-3p | -0.378 | 0.308578688 |
| hsa-miR-181a-2-3p | -0.325 | 0.316766943 |
| hsa-miR-203a-5p | -0.384 | 0.321786699 |
| hsa-miR-563 | 0.709 | 0.321786699 |
| hsa-miR-499a-5p | -0.455 | 0.325093559 |
| hsa-miR-652-3p | -0.625 | 0.325093559 |
| hsa-miR-922 | -0.556 | 0.325093559 |
| hsa-miR-7-5p | -0.605 | 0.325093559 |
| hsa-miR-137 | 0.481 | 0.325093559 |
| hsa-miR-181a-3p | -0.171 | 0.330192964 |
| hsa-miR-485-3p | -0.445 | 0.333558407 |
| hsa-miR-376a-3p | -0.933 | 0.333558407 |
| hsa-miR-487a-3p | 0.524 | 0.333558407 |
| hsa-miR-1283 | 0.942 | 0.333558407 |
| hsa-miR-125b-5p | -0.346 | 0.338755222 |
| hsa-miR-514b-5p | 0.173 | 0.342161278 |
| hsa-miR-409-3p | -0.453 | 0.350901883 |
| hsa-miR-376c-3p | -0.900 | 0.350901883 |
| hsa-miR-517c-3p+hsa-miR-519a-3p | -0.165 | 0.35628744 |
| hsa-miR-548m | -0.354 | 0.359779852 |
| hsa-miR-548aa+hsa-miR-548t-3p | 0.681 | 0.359779852 |
| hsa-miR-155-5p | -0.317 | 0.36879473 |
| hsa-miR-450a-5p | -0.570 | 0.37794598 |
| hsa-miR-3605-5p | -0.332 | 0.383637056 |
| hsa-miR-4488 | 0.413 | 0.393031943 |
| hsa-miR-508-5p | -0.324 | 0.393031943 |
| hsa-let-7c-5p | -0.573 | 0.396655021 |
| hsa-miR-136-5p | -0.345 | 0.396655021 |
| hsa-miR-23b-3p | 0.442 | 0.415900971 |
| hsa-miR-34a-5p | -0.520 | 0.415900971 |
| hsa-miR-1295a | -0.289 | 0.415900971 |
| hsa-miR-208a-3p | -0.091 | 0.422041825 |
| hsa-miR-3144-3p | -0.246 | 0.422041825 |
| hsa-miR-148a-3p | 0.266 | 0.425723035 |
| hsa-miR-29a-3p | 0.653 | 0.425723035 |
| hsa-miR-150-5p | -0.421 | 0.431983908 |
| hsa-miR-299-3p | 0.366 | 0.431983908 |
| hsa-miR-128-3p | -0.509 | 0.435676451 |
| hsa-miR-143-3p | -0.572 | 0.435676451 |
| hsa-miR-107 | 0.367 | 0.435676451 |
| hsa-miR-324-3p | 0.397 | 0.435676451 |
| hsa-miR-221-3p | 0.357 | 0.445760079 |
| hsa-miR-651-5p | -0.226 | 0.445760079 |
| hsa-miR-361-5p | 0.574 | 0.447148589 |
| hsa-let-7d-5p | 0.605 | 0.452269976 |
| hsa-miR-526b-5p | -0.117 | 0.452269976 |
| hsa-miR-18a-5p | 0.423 | 0.455972693 |
| hsa-miR-448 | 0.456 | 0.455972693 |
| hsa-miR-639 | 0.434 | 0.455972693 |
| hsa-miR-127-3p | -0.426 | 0.455972693 |
| hsa-miR-29c-3p | 0.630 | 0.466312978 |
| hsa-miR-337-3p | 0.273 | 0.466312978 |
| hsa-miR-1323 | 0.403 | 0.466312978 |
| hsa-miR-27b-3p | -0.014 | 0.467831374 |
| hsa-miR-19b-3p | -0.195 | 0.47308368 |
| hsa-miR-1185-2-3p | 0.455 | 0.476779535 |
| hsa-miR-627-3p | -0.016 | 0.48368496 |
| hsa-miR-93-5p | -0.022 | 0.487370875 |
| hsa-miR-30e-5p | 0.329 | 0.487370875 |
| hsa-miR-3613-3p | -0.129 | 0.494407197 |
| hsa-miR-10a-5p | -0.192 | 0.494413837 |
| hsa-miR-324-5p | -0.432 | 0.498085423 |
| hsa-miR-518b | -0.485 | 0.498085423 |
| hsa-miR-503-5p | -0.770 | 0.498085423 |
| hsa-miR-197-3p | 0.326 | 0.498085423 |
| hsa-miR-769-3p | -0.273 | 0.498085423 |
| hsa-miR-873-3p | -0.294 | 0.498085423 |
| hsa-miR-495-5p | 0.294 | 0.498085423 |
| hsa-miR-548q | -0.092 | 0.50526213 |
| hsa-miR-519c-3p | -0.273 | 0.505268679 |
| hsa-miR-1537-3p | -0.555 | 0.519877416 |
| hsa-miR-584-3p | -0.296 | 0.519877416 |
| hsa-miR-1278 | -0.191 | 0.527349264 |
| hsa-miR-342-3p | -0.620 | 0.530951282 |
| hsa-miR-650 | -0.191 | 0.53857128 |
| hsa-miR-335-5p | -0.563 | 0.542141201 |
| hsa-miR-191-5p | 0.514 | 0.542141201 |
| hsa-miR-199b-5p | -0.107 | 0.542141201 |
| hsa-miR-1290 | 0.092 | 0.544214473 |
| hsa-miR-337-5p | 0.393 | 0.549911806 |
| hsa-miR-942-5p | 0.014 | 0.561368749 |
| hsa-miR-301a-3p | 0.028 | 0.564861113 |
| hsa-miR-412-3p | -0.198 | 0.572934027 |
| hsa-miR-27a-3p | 0.039 | 0.572939929 |
| hsa-let-7b-5p | -0.347 | 0.576386857 |
| hsa-miR-4787-5p | -0.085 | 0.576386857 |
| hsa-miR-33b-5p | 0.415 | 0.578767651 |
| hsa-miR-377-3p | -0.118 | 0.588020158 |
| hsa-miR-543 | -0.653 | 0.596398879 |
| hsa-miR-1249-3p | -0.054 | 0.596410178 |
| hsa-miR-548ar-3p | -0.109 | 0.596415826 |
| hsa-miR-185-5p | -0.039 | 0.599758691 |
| hsa-miR-98-5p | 0.409 | 0.599758691 |
| hsa-miR-145-5p | -0.217 | 0.599758691 |
| hsa-miR-522-3p | 0.337 | 0.608315741 |
| hsa-miR-1224-3p | 0.003 | 0.608315741 |
| hsa-miR-134-3p | -0.054 | 0.608315741 |
| hsa-miR-378f | -0.415 | 0.611600049 |
| hsa-miR-3614-5p | 0.229 | 0.614305096 |
| hsa-miR-181d-3p | -0.321 | 0.620320288 |
| hsa-miR-425-5p | 0.347 | 0.62354175 |
| hsa-miR-20a-5p+hsa-miR-20b-5p | -0.385 | 0.62354175 |
| hsa-miR-370-3p | -0.354 | 0.62354175 |
| hsa-miR-197-5p | -0.373 | 0.626360985 |
| hsa-miR-28-5p | -0.468 | 0.632426854 |
| hsa-miR-199a-5p | 0.045 | 0.638517555 |
| hsa-miR-106a-5p+hsa-miR-17-5p | -0.252 | 0.644632744 |
| hsa-miR-187-3p | 0.196 | 0.644632744 |
| hsa-miR-4536-3p | -0.098 | 0.644632744 |
| hsa-miR-625-5p | -0.093 | 0.647715879 |
| hsa-miR-570-3p | 0.475 | 0.647715879 |
| hsa-miR-19a-3p | -0.244 | 0.659942969 |
| hsa-miR-374a-5p | 0.467 | 0.659942969 |
| hsa-miR-937-3p | -0.227 | 0.659942969 |
| hsa-miR-382-5p | -0.384 | 0.672259731 |
| hsa-miR-585-3p | 0.004 | 0.675563603 |
| hsa-miR-615-3p | -0.173 | 0.681813584 |
| hsa-miR-576-3p | 0.024 | 0.68181821 |
| hsa-miR-222-3p | 0.300 | 0.68466332 |
| hsa-miR-548a-5p | 0.169 | 0.68466332 |
| hsa-miR-4707-5p | 0.033 | 0.697150823 |
| hsa-miR-18b-5p | -0.006 | 0.7070479 |
| hsa-miR-5001-5p | 0.130 | 0.707052198 |
| hsa-let-7f-5p | 0.089 | 0.709719264 |
| hsa-miR-660-5p | 0.139 | 0.709719264 |
| hsa-miR-208b-3p | -0.092 | 0.719793063 |
| hsa-miR-126-3p | 0.135 | 0.7223656 |
| hsa-miR-642a-5p | 0.156 | 0.7223656 |
| hsa-miR-411-5p | 0.168 | 0.7223656 |
| hsa-miR-95-3p | 0.194 | 0.73260829 |
| hsa-miR-146a-5p | 0.086 | 0.73508673 |
| hsa-miR-612 | -0.426 | 0.73508673 |
| hsa-miR-130b-3p | -0.031 | 0.74550646 |
| hsa-miR-363-3p | -0.165 | 0.747879492 |
| hsa-miR-1296-3p | 0.330 | 0.747879492 |
| hsa-miR-4536-5p | -0.143 | 0.760740669 |
| hsa-miR-513a-3p | 0.175 | 0.760740669 |
| hsa-miR-582-5p | -0.055 | 0.771503135 |
| hsa-miR-664a-3p | 0.223 | 0.771506552 |
| hsa-miR-210-5p | 0.030 | 0.771506552 |
| hsa-miR-598-3p | 0.056 | 0.773666987 |
| hsa-miR-200a-3p | -0.293 | 0.773666987 |
| hsa-miR-512-5p | 0.353 | 0.773666987 |
| hsa-miR-1185-5p | -0.118 | 0.773666987 |
| hsa-miR-148b-3p | -0.274 | 0.786655124 |
| hsa-miR-25-3p | -0.072 | 0.786655124 |
| hsa-miR-125a-5p | 0.070 | 0.786655124 |
| hsa-miR-1197 | -0.106 | 0.786655124 |
| hsa-miR-210-3p | 0.220 | 0.797765869 |
| hsa-miR-525-5p | 0.049 | 0.797765869 |
| hsa-miR-487b-3p | -0.099 | 0.799701705 |
| hsa-miR-378i | 0.064 | 0.799701705 |
| hsa-miR-30b-5p | -0.100 | 0.799701705 |
| hsa-miR-302d-3p | -0.118 | 0.804367898 |
| hsa-miR-302a-3p | 0.046 | 0.81098391 |
| hsa-miR-3168 | -0.074 | 0.812803311 |
| hsa-miR-26b-5p | -0.001 | 0.812803311 |
| hsa-miR-603 | -0.091 | 0.812803311 |
| hsa-miR-378d | -0.306 | 0.812803311 |
| hsa-miR-133a-3p | 0.199 | 0.824256061 |
| hsa-miR-548g-3p | 0.139 | 0.825956476 |
| hsa-miR-374b-5p | -0.033 | 0.825956476 |
| hsa-miR-574-3p | -0.083 | 0.825956476 |
| hsa-miR-186-5p | -0.379 | 0.825956476 |
| hsa-miR-211-5p | 0.161 | 0.837578655 |
| hsa-miR-4284 | -0.099 | 0.837578655 |
| hsa-miR-933 | 0.047 | 0.837578655 |
| hsa-miR-32-5p | -0.135 | 0.839157696 |
| hsa-miR-299-5p | 0.320 | 0.839157696 |
| hsa-miR-4485-3p | 0.218 | 0.850943452 |
| hsa-miR-21-5p | 0.049 | 0.852403424 |
| hsa-miR-15a-5p | 0.260 | 0.852403424 |
| hsa-let-7e-5p | 0.063 | 0.852403424 |
| hsa-miR-223-3p | -0.239 | 0.864360295 |
| hsa-miR-371a-5p | 0.363 | 0.864360295 |
| hsa-miR-487b-5p | -0.048 | 0.864360295 |
| hsa-miR-30e-3p | 0.029 | 0.865690082 |
| hsa-miR-421 | -0.096 | 0.865690082 |
| hsa-miR-551b-3p | 0.183 | 0.865690082 |
| hsa-miR-378h | -0.029 | 0.865690082 |
| hsa-miR-369-3p | -0.038 | 0.877809937 |
| hsa-miR-514a-3p | 0.062 | 0.877811802 |
| hsa-miR-199a-3p+hsa-miR-199b-3p | -0.141 | 0.879014054 |
| hsa-miR-28-3p | -0.114 | 0.879014054 |
| hsa-miR-301a-5p | 0.157 | 0.879014054 |
| hsa-miR-365a-3p+hsa-miR-365b-3p | -0.197 | 0.879014054 |
| hsa-miR-92a-3p | 0.074 | 0.884551061 |
| hsa-miR-181a-5p | 0.128 | 0.884551061 |
| hsa-miR-5010-3p | 0.260 | 0.891298683 |
| hsa-miR-140-5p | -0.277 | 0.892371696 |
| hsa-miR-630 | 0.211 | 0.892371696 |
| hsa-miR-891a-5p | -0.023 | 0.892371696 |
| hsa-miR-626 | 0.064 | 0.904817085 |
| hsa-miR-328-5p | -0.035 | 0.904817085 |
| hsa-miR-106b-5p | -0.187 | 0.905759337 |
| hsa-miR-548ar-5p | -0.088 | 0.905759337 |
| hsa-miR-548j-3p | 0.202 | 0.905759337 |
| hsa-miR-381-3p | 0.102 | 0.91917328 |
| hsa-miR-542-3p | 0.019 | 0.931932912 |
| hsa-miR-1268b | -0.032 | 0.945522508 |
| hsa-miR-23a-3p | -0.107 | 0.946065187 |
| hsa-miR-122-5p | 0.060 | 0.946065187 |
| hsa-miR-423-5p | 0.018 | 0.946065187 |
| hsa-miR-10b-5p | 0.007 | 0.946065187 |
| hsa-miR-15b-5p | -0.042 | 0.946065187 |
| hsa-miR-378g | 0.000 | 0.946065187 |
| hsa-miR-301b-5p | -0.071 | 0.946065187 |
| hsa-miR-423-3p | 0.010 | 0.959127975 |
| hsa-miR-4647 | 0.090 | 0.959127975 |
| hsa-miR-593-3p | 0.048 | 0.959127975 |
| hsa-miR-30d-5p | -0.042 | 0.959535664 |
| hsa-miR-766-3p | -0.063 | 0.959535664 |
| hsa-miR-1302 | 0.039 | 0.959535664 |
| hsa-miR-510-3p | 0.047 | 0.973017474 |
| hsa-miR-767-5p | -0.005 | 0.973017474 |
| hsa-miR-498 | -0.088 | 0.986370691 |
| hsa-miR-597-5p | -0.065 | 0.986370691 |
| hsa-miR-454-3p | -0.068 | 0.986370691 |
| hsa-miR-1262 | -0.021 | 0.986370691 |
| hsa-miR-16-5p | -0.171 | 0.986506846 |
| hsa-miR-379-5p | -0.191 | 1 |
| hsa-miR-182-3p | 0.076 | 1 |
| hsa-miR-497-5p | -0.121 | 1 |
| [illegible] | [illegible] | [illegible] |

**All samples**

| miRNA | BH adjusted MW test P value - Mild vs | LFC |
|---|---|---|
| hsa-miR-451a | 0.045920855 | 0.926 |
| hsa-miR-495-3p | 0.045920855 | 1.475 |
| hsa-miR-376a-3p | 0.072876128 | 1.178 |
| hsa-miR-340-5p | 0.072876128 | 0.899 |
| hsa-miR-148b-3p | 0.072876128 | 0.745 |
| hsa-let-7b-5p | 0.072876128 | 0.906 |
| hsa-miR-146a-5p | 0.072876128 | 1.136 |
| hsa-miR-423-5p | 0.072876128 | 0.712 |
| hsa-miR-23a-3p | 0.072876128 | 0.857 |
| hsa-miR-432-5p | 0.093654396 | 1.256 |
| hsa-miR-499a-5p | 0.096529768 | 0.826 |
| hsa-miR-150-5p | 0.096529768 | 1.403 |
| hsa-miR-223-3p | 0.117156603 | 0.839 |
| hsa-miR-199a-3p | 0.117234382 | 0.975 |
| hsa-miR-21-5p | 0.117848134 | 0.496 |
| hsa-miR-143-3p | 0.119513373 | 0.958 |
| hsa-miR-122-5p | 0.12763151 | 1.382 |
| hsa-miR-543 | 0.129978442 | 0.924 |
| hsa-miR-1537-3p | 0.129978442 | 1.061 |
| hsa-miR-10b-5p | 0.136153874 | 0.852 |
| hsa-miR-25-3p | 0.136153874 | 0.669 |
| hsa-miR-433-3p | 0.136821842 | 0.933 |
| hsa-miR-106a-5p | 0.145590997 | 0.876 |
| hsa-miR-382-5p | 0.150175356 | 1.302 |
| hsa-miR-185-5p | 0.150175356 | 0.700 |
| hsa-miR-323a-3p | 0.167506797 | 0.894 |
| hsa-miR-188-5p | 0.167506797 | 0.583 |
| hsa-miR-376c-3p | 0.167506797 | 1.193 |
| hsa-miR-142-3p | 0.167506797 | 0.558 |
| hsa-miR-191-5p | 0.167506797 | 1.178 |
| hsa-miR-16-5p | 0.167506797 | 0.511 |
| hsa-miR-186-5p | 0.167506797 | 0.622 |
| hsa-miR-625-5p | 0.167506797 | 0.680 |
| hsa-miR-26b-5p | 0.167506797 | 0.745 |
| hsa-miR-485-3p | 0.179904189 | 0.752 |
| hsa-miR-92a-3p | 0.179904189 | 0.754 |
| hsa-miR-199a-5p | 0.191280015 | 0.703 |
| hsa-miR-152-3p | 0.194210605 | 0.545 |
| hsa-miR-4461 | 0.206340256 | 0.530 |
| hsa-miR-19a-3p | 0.206340256 | 0.877 |
| hsa-miR-20a-5p+h | 0.208023313 | 0.806 |
| hsa-miR-301b-3p | 0.210854977 | 0.684 |
| hsa-miR-126-3p | 0.210854977 | 0.613 |
| hsa-miR-19b-3p | 0.224273639 | 0.787 |
| hsa-miR-134-3p | 0.227321497 | 0.544 |
| hsa-let-7g-5p | 0.22775457 | 0.628 |
| hsa-miR-4536-3p | 0.238064659 | 0.643 |
| hsa-miR-93-5p | 0.238064659 | 0.977 |
| hsa-miR-4536-5p | 0.238582255 | 0.614 |
| hsa-miR-128-3p | 0.238582255 | 0.775 |
| hsa-miR-922 | 0.241333528 | 0.575 |
| hsa-miR-409-3p | 0.241333528 | 0.749 |
| hsa-miR-342-3p | 0.241333528 | 0.840 |
| hsa-miR-1278 | 0.24259251 | 0.469 |
| hsa-miR-381-3p | 0.24259251 | 0.545 |
| hsa-let-7c-5p | 0.274069147 | 0.922 |
| hsa-miR-107 | 0.282629044 | 0.813 |
| hsa-miR-361-5p | 0.282949265 | 0.786 |
| hsa-miR-302d-3p | 0.29828323 | 0.463 |
| hsa-let-7d-5p | 0.299802904 | 0.840 |
| hsa-miR-4707-5p | 0.309125297 | 0.485 |
| hsa-miR-942-5p | 0.314155226 | 0.556 |
| hsa-miR-627-3p | 0.317285395 | 0.506 |
| hsa-miR-1246 | 0.317285395 | 0.576 |
| hsa-miR-548g-3p | 0.317285395 | 0.676 |
| hsa-miR-181d-3p | 0.321764813 | 0.432 |
| hsa-miR-328-5p | 0.321764813 | 0.498 |
| hsa-miR-7-5p | 0.321764813 | 0.554 |
| hsa-miR-518b | 0.321764813 | 0.649 |
| hsa-miR-98-5p | 0.321764813 | 0.810 |
| hsa-miR-212-3p | 0.333766139 | 0.428 |
| hsa-miR-144-3p | 0.333766139 | 0.847 |
| hsa-miR-548m | 0.333766139 | 0.476 |
| hsa-miR-873-3p | 0.333766139 | 0.462 |
| hsa-miR-5001-5p | 0.333766139 | 0.454 |
| hsa-miR-15a-5p | 0.333766139 | 0.902 |
| hsa-miR-15b-5p | 0.333766139 | 0.569 |
| hsa-miR-378i | 0.333766139 | 0.552 |
| hsa-miR-371a-5p | 0.338601892 | 0.411 |
| hsa-miR-22-3p | 0.341212039 | 0.939 |
| hsa-miR-208b-3p | 0.341563986 | 0.639 |
| hsa-miR-612 | 0.343103831 | 0.650 |
| hsa-miR-148a-3p | 0.349763378 | 0.300 |
| hsa-miR-132-3p | 0.349763378 | 0.547 |
| hsa-miR-26a-5p | 0.358454387 | 0.595 |
| hsa-miR-363-3p | 0.360157826 | 0.587 |
| hsa-miR-3144-3p | 0.360256894 | 0.381 |
| hsa-let-7a-5p | 0.360256894 | 0.351 |
| hsa-miR-210-5p | 0.360256894 | 0.437 |
| hsa-miR-1249-3p | 0.360256894 | 0.478 |
| hsa-miR-210-3p | 0.360256894 | 0.411 |
| hsa-miR-301a-5p | 0.360256894 | 0.552 |
| hsa-miR-1973 | 0.360256894 | 0.609 |
| hsa-miR-548a-5p | 0.360256894 | 0.504 |
| hsa-miR-181a-5p | 0.360256894 | 0.805 |
| hsa-miR-151a-5p | 0.360256894 | 0.727 |
| hsa-miR-505-3p | 0.369289982 | 0.387 |
| hsa-miR-651-5p | 0.375845422 | 0.387 |
| hsa-miR-181a-3p | 0.375845422 | 0.420 |
| hsa-miR-508-5p | 0.386150325 | 0.465 |
| hsa-miR-27b-3p | 0.386150325 | 0.550 |
| hsa-miR-3613-3p | 0.393217607 | 0.478 |
| hsa-miR-519c-3p | 0.393792535 | 0.395 |
| hsa-miR-6721-5p | 0.393792535 | 0.381 |
| hsa-miR-370-3p | 0.393792535 | 0.543 |
| hsa-miR-378d | 0.393792535 | 0.548 |
| hsa-miR-514b-5p | 0.393792535 | 0.515 |
| hsa-miR-208a-3p | 0.402238781 | 0.539 |
| hsa-miR-130a-3p | 0.40340927 | 0.339 |
| hsa-miR-374a-5p | 0.40340927 | 0.646 |
| hsa-miR-517c-3p | 0.40340927 | 0.353 |
| hsa-miR-548v | 0.413350963 | 0.390 |
| hsa-miR-197-3p | 0.413350963 | 0.354 |
| hsa-miR-324-5p | 0.413350963 | 0.409 |
| hsa-miR-4516 | 0.41847288 | -0.393 |
| hsa-miR-30e-3p | 0.41847288 | 0.552 |
| hsa-miR-548ah-5p | 0.429948104 | 0.402 |
| hsa-miR-3605-5p | 0.429948104 | 0.440 |
| hsa-miR-378h | 0.432767178 | 0.452 |
| hsa-miR-30d-5p | 0.432767178 | 0.764 |
| hsa-miR-187-3p | 0.432946154 | 0.520 |
| hsa-miR-4787-5p | 0.437138597 | 0.394 |
| hsa-miR-526b-5p | 0.437138597 | 0.325 |
| hsa-miR-221-3p | 0.455284981 | 0.624 |
| hsa-miR-377-3p | 0.455284981 | 0.368 |
| hsa-miR-585-3p | 0.455284981 | 0.412 |
| hsa-miR-425-5p | 0.455284981 | 0.631 |
| hsa-miR-299-3p | 0.455284981 | 0.486 |
| hsa-miR-133a-3p | 0.455284981 | 0.435 |
| hsa-miR-1224-3p | 0.455607814 | 0.290 |
| hsa-miR-411-5p | 0.458865939 | 0.497 |
| hsa-miR-32-5p | 0.458865939 | 0.378 |
| hsa-miR-548ar-3p | 0.46475907 | 0.450 |
| hsa-miR-503-5p | 0.46475907 | 0.608 |
| hsa-miR-769-3p | 0.475919691 | 0.308 |
| hsa-miR-27a-3p | 0.475919691 | 0.414 |
| hsa-miR-576-3p | 0.476397576 | 0.381 |
| hsa-miR-1295a | 0.476397576 | 0.481 |
| hsa-miR-532-3p | 0.476397576 | 0.416 |
| hsa-miR-337-3p | 0.476397576 | 0.415 |
| hsa-miR-548j-3p | 0.487291213 | 0.294 |
| hsa-miR-369-3p | 0.489084921 | 0.479 |
| hsa-miR-4485-3p | 0.498506866 | 0.298 |
| hsa-miR-582-5p | 0.498506866 | 0.357 |
| hsa-miR-1255a | 0.498506866 | 0.366 |
| hsa-miR-525-5p | 0.499406143 | 0.402 |
| hsa-miR-603 | 0.499406143 | 0.365 |
| hsa-miR-29c-3p | 0.499406143 | 0.566 |
| hsa-miR-454-3p | 0.499406143 | 0.297 |
| hsa-miR-764 | 0.505252503 | 0.247 |
| hsa-miR-769-5p | 0.505766531 | 0.251 |
| hsa-miR-10a-5p | 0.505766531 | 0.345 |
| hsa-miR-487a-3p | 0.505766531 | 0.211 |
| hsa-miR-203a-5p | 0.506745766 | 0.311 |
| hsa-miR-548ar-5p | 0.506745766 | 0.297 |
| hsa-miR-412-3p | 0.506745766 | 0.284 |
| hsa-miR-301b-5p | 0.506745766 | 0.354 |
| hsa-miR-182-3p | 0.506745766 | 0.334 |
| hsa-miR-644a | 0.506745766 | 0.334 |
| hsa-miR-28-5p | 0.506745766 | 0.354 |
| hsa-miR-337-5p | 0.506745766 | 0.404 |
| hsa-miR-28-3p | 0.506745766 | 0.403 |
| hsa-miR-937-3p | 0.512115869 | 0.403 |
| hsa-miR-301a-3p | 0.512115869 | 0.466 |
| hsa-miR-1228-3p | 0.520670361 | 0.340 |
| hsa-miR-4284 | 0.524817755 | 0.306 |
| hsa-miR-34a-5p | 0.524817755 | 0.342 |
| hsa-miR-590-5p | 0.528214535 | 0.318 |
| hsa-miR-1972 | 0.538999498 | -0.710 |
| hsa-miR-335-5p | 0.538999498 | 0.529 |
| hsa-miR-183-5p | 0.539797603 | 0.354 |
| hsa-miR-630 | 0.543245637 | -0.466 |
| hsa-miR-374b-5p | 0.546706375 | 0.045 |
| hsa-miR-30e-5p | 0.548851774 | 0.426 |
| hsa-miR-650 | 0.550514984 | 0.297 |
| hsa-miR-522-3p | 0.550514984 | 0.451 |
| hsa-miR-137 | 0.558780406 | 0.322 |
| hsa-miR-3147 | 0.558780406 | 0.356 |
| hsa-miR-450a-5p | 0.558780406 | 0.285 |
| hsa-miR-570-3p | 0.558780406 | 0.285 |
| hsa-miR-378g | 0.558780406 | 0.348 |
| hsa-miR-18a-5p | 0.562308343 | 0.407 |
| hsa-miR-302a-3p | 0.571166078 | 0.345 |
| hsa-miR-379-5p | 0.572053298 | 0.251 |
| hsa-miR-497-5p | 0.586355611 | -0.274 |
| hsa-miR-652-3p | 0.588590457 | 0.188 |
| hsa-miR-18b-5p | 0.589480912 | 0.196 |
| hsa-miR-125b-5p | 0.593544274 | 0.209 |
| hsa-miR-597-5p | 0.593544274 | 0.384 |
| hsa-miR-197-5p | 0.593544274 | 0.302 |
| hsa-miR-514a-3p | 0.618956002 | 0.222 |
| hsa-miR-33b-5p | 0.622604846 | 0.336 |
| hsa-miR-127-3p | 0.631795418 | 0.304 |
| hsa-miR-626 | 0.634073876 | 0.316 |
| hsa-miR-324-3p | 0.634971575 | 0.248 |
| hsa-miR-1285-5p | 0.640033547 | -0.233 |
| hsa-miR-548aa+h | 0.640480965 | 0.102 |
| hsa-miR-222-3p | 0.640480965 | 0.211 |
| hsa-miR-542-3p | 0.640480965 | 0.490 |
| hsa-miR-365a-3p | 0.640480965 | 0.380 |
| hsa-miR-30b-5p | 0.640480965 | 0.056 |
| hsa-miR-130b-3p | 0.653834707 | 0.161 |
| hsa-miR-423-3p | 0.660314059 | 0.248 |
| hsa-miR-3168 | 0.666807225 | 0.236 |
| hsa-miR-106b-5p | 0.671439614 | 0.473 |
| hsa-miR-642a-5p | 0.671439614 | 0.228 |
| hsa-miR-151a-3p | 0.675153075 | 0.196 |
| hsa-miR-448 | 0.692404794 | 0.236 |
| hsa-miR-3614-5p | 0.692404794 | 0.162 |
| hsa-let-7f-5p | 0.718847787 | -0.327 |
| hsa-miR-487b-3p | 0.724013481 | 0.265 |
| hsa-miR-487b-5p | 0.756043998 | 0.210 |
| hsa-miR-378e | 0.756043998 | 0.251 |
| hsa-miR-1185-5p | 0.764149296 | 0.247 |
| hsa-miR-362-5p | 0.772181638 | 0.190 |
| hsa-miR-1290 | 0.772181638 | 0.161 |
| hsa-miR-145-5p | 0.772181638 | 0.122 |
| hsa-miR-378f | 0.772181638 | 0.198 |
| hsa-miR-563 | 0.772181638 | 0.121 |
| hsa-miR-1305 | 0.772181638 | 0.242 |
| hsa-miR-627-5p | 0.772181638 | 0.204 |
| hsa-miR-450a-2-3 | 0.772181638 | 0.225 |
| hsa-miR-4454+hs | 0.789248288 | -0.106 |
| hsa-miR-23b-3p | 0.789248288 | 0.084 |
| hsa-miR-767-5p | 0.789248288 | 0.191 |
| hsa-miR-513a-3p | 0.789248288 | 0.258 |
| hsa-miR-4286 | 0.789248288 | 0.215 |
| hsa-miR-136-5p | 0.79089871 | 0.196 |
| hsa-miR-574-3p | 0.79089871 | 0.068 |
| hsa-miR-551b-3p | 0.803341565 | 0.062 |
| hsa-miR-933 | 0.803341565 | 0.234 |
| hsa-let-7e-5p | 0.803341565 | 0.020 |
| hsa-miR-639 | 0.816991296 | 0.224 |
| hsa-miR-1253 | 0.818310423 | 0.057 |
| hsa-miR-939-5p | 0.818310423 | 0.268 |
| hsa-miR-125a-5p | 0.818310423 | -0.086 |
| hsa-miR-4488 | 0.818310423 | 0.185 |
| hsa-miR-5010-3p | 0.818310423 | 0.240 |
| hsa-miR-891a-5p | 0.818310423 | -0.009 |
| hsa-miR-766-3p | 0.818310423 | 0.029 |
| hsa-miR-95-3p | 0.820519203 | -0.021 |
| hsa-miR-200a-3p | 0.825722281 | 0.253 |
| hsa-miR-140-5p | 0.825722281 | -0.061 |
| hsa-miR-181a-2-3 | 0.825722281 | -0.119 |
| hsa-miR-299-5p | 0.839077674 | 0.158 |
| hsa-miR-574-5p | 0.84823253 | -0.159 |
| hsa-miR-548q | 0.850379741 | 0.087 |
| hsa-miR-1283 | 0.852521478 | 0.302 |
| hsa-miR-29b-3p | 0.852624656 | 0.141 |
| hsa-miR-1323 | 0.852624656 | 0.156 |
| hsa-miR-1262 | 0.86584593 | 0.204 |
| hsa-miR-4647 | 0.870720736 | 0.119 |
| hsa-miR-593-3p | 0.889460547 | 0.082 |
| hsa-miR-660-5p | 0.910985201 | 0.112 |
| hsa-miR-584-3p | 0.911583575 | 0.127 |
| hsa-miR-615-3p | 0.913972875 | 0.142 |
| hsa-miR-1302 | 0.913972875 | 0.155 |
| hsa-miR-510-3p | 0.913972875 | 0.132 |
| hsa-miR-802 | 0.913972875 | -0.078 |
| hsa-miR-495-5p | 0.924176054 | 0.097 |
| hsa-miR-211-5p | 0.94242397 | 0.044 |
| hsa-miR-25-5p | 0.94242397 | 0.090 |
| hsa-miR-155-5p | 0.94242397 | 0.092 |
| hsa-miR-502-5p | 0.94242397 | 0.066 |
| hsa-miR-199b-5p | 0.94242397 | 0.035 |
| hsa-miR-664a-3p | 0.94242397 | 0.122 |
| hsa-miR-421 | 0.948335068 | 0.081 |
| hsa-miR-184 | 0.950176884 | 0.017 |
| hsa-miR-1268b | 0.950667201 | 0.038 |
| hsa-miR-598-3p | 0.963198804 | 0.078 |
| hsa-miR-548e-5p | 0.964982008 | 0.106 |
| hsa-miR-24-3p | 0.965865575 | -0.025 |
| hsa-miR-498 | 0.965865575 | 0.075 |
| hsa-miR-1197 | 0.978204459 | -0.047 |
| hsa-miR-320e | 0.978997037 | 0.033 |
| hsa-miR-888-5p | 0.978997037 | 0.015 |
| hsa-miR-29a-3p | 0.984628379 | -0.023 |
| hsa-miR-512-5p | 0.985372701 | 0.038 |
| hsa-miR-1185-2-3 | 0.985372701 | 0.140 |
| hsa-miR-1296-3p | 0.989629983 | 0.043 |

**First samples only**

| miRNA | MW test P value Mild vs | LFC Mild- |
|---|---|---|
| hsa-miR-590-5p | 0.010246119 | 1.527 |
| hsa-miR-1973 | 0.011296875 | 1.397 |
| hsa-miR-548g-3p | 0.018076821 | 1.710 |
| hsa-miR-128-3p | 0.020695546 | 1.445 |
| hsa-miR-495-3p | 0.02163768 | 1.298 |
| hsa-miR-143-3p | 0.02468684 | 1.327 |
| hsa-miR-151a-5p | 0.026916461 | 1.290 |
| hsa-miR-499a-5p | 0.028093793 | 1.013 |
| hsa-miR-485-3p | 0.030579466 | 1.403 |
| hsa-miR-340-5p | 0.031409001 | 0.629 |
| hsa-miR-363-3p | 0.034653949 | 1.473 |
| hsa-miR-150-5p | 0.034737619 | 1.672 |
| hsa-miR-324-5p | 0.036109413 | 1.391 |
| hsa-miR-188-5p | 0.040785892 | 0.617 |
| hsa-miR-518b | 0.040785892 | 1.498 |
| hsa-miR-323a-3p | 0.044175288 | 1.254 |
| hsa-miR-144-3p | 0.045955587 | 0.853 |
| hsa-miR-1537-3p | 0.045955587 | 1.355 |
| hsa-let-7b-5p | 0.045955587 | 0.715 |
| hsa-miR-19a-3p | 0.051655558 | 0.941 |
| hsa-miR-342-3p | 0.057923863 | 1.266 |
| hsa-miR-432-5p | 0.062797288 | 1.133 |
| hsa-miR-451a | 0.066305166 | 0.723 |
| hsa-let-7c-5p | 0.07232158 | 1.184 |
| hsa-miR-652-3p | 0.077715332 | 0.986 |
| hsa-miR-409-3p | 0.080530336 | 1.220 |
| hsa-miR-625-5p | 0.080530336 | 0.938 |
| hsa-miR-23a-3p | 0.080530336 | 0.545 |
| hsa-miR-433-3p | 0.083426067 | 0.892 |
| hsa-miR-3168 | 0.083426067 | 0.884 |
| hsa-miR-28-5p | 0.089351098 | 1.118 |
| hsa-miR-152-3p | 0.10054795 | 0.609 |
| hsa-miR-505-3p | 0.106080293 | 0.684 |
| hsa-miR-644a | 0.106080293 | 0.568 |
| hsa-miR-376a-3p | 0.106080293 | 0.945 |
| hsa-miR-337-5p | 0.107431856 | 0.935 |
| hsa-miR-151a-3p | 0.109674692 | 0.893 |
| hsa-miR-6721-5p | 0.118451507 | 0.674 |
| hsa-miR-106a-5p+hsa-miR-17-5p | 0.120371399 | 0.813 |
| hsa-miR-199a-3p+hsa-miR-199b-3p | 0.121026458 | 0.726 |
| hsa-miR-223-3p | 0.124284188 | 0.762 |
| hsa-miR-107 | 0.125004538 | 0.795 |
| hsa-miR-320e | 0.128295498 | -0.484 |
| hsa-miR-185-5p | 0.129081973 | 0.644 |
| hsa-miR-425-5p | 0.1332601 | 0.905 |
| hsa-miR-19b-3p | 0.13450026 | 0.917 |
| hsa-miR-148b-3p | 0.141923673 | 0.458 |
| hsa-miR-26b-5p | 0.141923673 | 0.568 |
| hsa-miR-381-3p | 0.141923673 | 0.691 |
| hsa-miR-423-3p | 0.143131055 | 0.849 |
| hsa-miR-1285-5p | 0.146411685 | -0.606 |
| hsa-miR-199a-5p | 0.147602821 | 0.561 |
| hsa-miR-92a-3p | 0.149878373 | 0.567 |
| hsa-miR-543 | 0.1544906 | 0.866 |
| hsa-miR-30e-3p | 0.155706385 | 0.940 |
| hsa-miR-122-5p | 0.155706385 | 1.132 |
| hsa-miR-30d-5p | 0.155706385 | 0.577 |
| hsa-miR-335-5p | 0.160515482 | 0.996 |
| hsa-miR-26a-5p | 0.170462971 | 0.534 |
| hsa-miR-548m | 0.170462971 | 0.546 |
| hsa-miR-450a-5p | 0.170462971 | 0.844 |
| hsa-miR-21-5p | 0.170462971 | 0.401 |
| hsa-miR-382-5p | 0.175603589 | 1.068 |
| hsa-miR-222-3p | 0.175603589 | 0.864 |
| hsa-miR-517c-3p+hsa-miR-519a-3p | 0.17670349 | 0.537 |
| hsa-miR-664a-3p | 0.17670349 | 0.765 |
| hsa-miR-423-5p | 0.18622388 | 0.494 |
| hsa-miR-20a-5p+hsa-miR-20b-5p | 0.191705569 | 0.871 |
| hsa-miR-374a-5p | 0.191705569 | 1.195 |
| hsa-miR-146a-5p | 0.191705569 | 0.598 |
| hsa-miR-10b-5p | 0.191705569 | 0.766 |
| hsa-miR-15b-5p | 0.191705569 | 0.749 |
| hsa-miR-582-5p | 0.192760256 | 0.632 |
| hsa-miR-3147 | 0.1927671 | 0.609 |
| hsa-miR-29c-3p | 0.197302899 | 0.962 |
| hsa-miR-487b-3p | 0.197302899 | 0.735 |
| hsa-miR-4461 | 0.198353214 | 0.321 |
| hsa-miR-93-5p | 0.208848081 | 0.762 |
| hsa-miR-16-5p | 0.208848081 | 0.398 |
| hsa-miR-208b-3p | 0.209878788 | 0.578 |
| hsa-let-7d-5p | 0.212834473 | 0.952 |
| hsa-miR-132-3p | 0.214797624 | 0.686 |
| hsa-miR-126-3p | 0.214797624 | 0.481 |
| hsa-miR-548e-5p | 0.218835765 | -0.579 |
| hsa-miR-376c-3p | 0.22086619 | 0.835 |
| hsa-miR-133a-3p | 0.221881581 | 0.659 |
| hsa-miR-522-3p | 0.224957644 | 0.577 |
| hsa-miR-221-3p | 0.227054513 | 0.656 |
| hsa-miR-4707-5p | 0.233363279 | 0.486 |
| hsa-miR-764 | 0.237566264 | 0.468 |
| hsa-miR-18a-5p | 0.239793123 | 0.839 |
| hsa-miR-922 | 0.246344633 | 0.670 |
| hsa-let-7f-5p | 0.246344633 | 0.678 |
| hsa-miR-191-5p | 0.253018344 | 0.880 |
| hsa-miR-548ar-3p | 0.254018335 | 0.548 |
| hsa-miR-23b-3p | 0.259814737 | 0.582 |
| hsa-miR-448 | 0.259814737 | 0.543 |
| hsa-miR-25-3p | 0.259814737 | 0.445 |
| hsa-miR-1224-3p | 0.260816195 | 0.441 |
| hsa-miR-27a-3p | 0.264261897 | 0.670 |
| hsa-miR-22-3p | 0.266734239 | 0.543 |
| hsa-miR-374b-5p | 0.266734239 | 0.585 |
| hsa-miR-140-5p | 0.266734239 | 0.610 |
| hsa-miR-1972 | 0.273777224 | -0.866 |
| hsa-miR-142-3p | 0.273777224 | 0.312 |
| hsa-miR-660-5p | 0.273777224 | 0.488 |
| hsa-miR-574-3p | 0.273777224 | 0.481 |
| hsa-miR-32-5p | 0.273777224 | 0.795 |
| hsa-miR-1278 | 0.27478677 | 0.283 |
| hsa-miR-650 | 0.281960319 | 0.438 |
| hsa-miR-299-3p | 0.285594313 | 0.587 |
| hsa-miR-1268b | 0.285594313 | -0.303 |
| hsa-let-7g-5p | 0.28823484 | 0.424 |
| hsa-miR-1290 | 0.292942068 | -0.342 |
| hsa-miR-377-3p | 0.295649928 | 0.439 |
| hsa-miR-378i | 0.295649928 | 0.404 |
| hsa-miR-379-5p | 0.295649928 | 0.523 |
| hsa-miR-212-3p | 0.304237773 | 0.462 |
| hsa-miR-5001-5p | 0.304237773 | 0.346 |
| hsa-miR-182-3p | 0.304237773 | 0.471 |
| hsa-miR-802 | 0.310853353 | -0.572 |
| hsa-miR-7-5p | 0.326554643 | 0.686 |
| hsa-miR-503-5p | 0.326554643 | 0.905 |
| hsa-miR-98-5p | 0.326554643 | 0.876 |
| hsa-miR-361-5p | 0.327654081 | 0.252 |
| hsa-miR-130a-3p | 0.334591826 | 0.234 |
| hsa-miR-197-3p | 0.334591826 | 0.493 |
| hsa-miR-125b-5p | 0.335712806 | 0.351 |
| hsa-miR-498 | 0.339789615 | 0.579 |
| hsa-miR-15a-5p | 0.35103838 | 0.556 |
| hsa-miR-208a-3p | 0.352209556 | 0.440 |
| hsa-miR-106b-5p | 0.359447197 | 0.786 |
| hsa-miR-148a-3p | 0.367979229 | 0.479 |
| hsa-miR-28-3p | 0.376634029 | 0.400 |
| hsa-miR-378g | 0.376634029 | 0.509 |
| hsa-miR-766-3p | 0.376634029 | 0.347 |
| hsa-miR-27b-3p | 0.382290186 | 0.454 |
| hsa-miR-181a-5p | 0.382290186 | 0.671 |
| hsa-miR-369-3p | 0.38670518 | 0.506 |
| hsa-miR-548aa+hsa-miR-548t-3p | 0.394309815 | -0.477 |
| hsa-miR-29a-3p | 0.403329567 | 0.438 |
| hsa-miR-34a-5p | 0.412469619 | 0.510 |
| hsa-miR-421 | 0.412469619 | 0.533 |
| hsa-miR-615-3p | 0.413888877 | 0.322 |
| hsa-miR-526b-5p | 0.413896058 | 0.111 |
| hsa-miR-1295a | 0.421729181 | 0.381 |
| hsa-miR-598-3p | 0.421729181 | 0.569 |
| hsa-miR-181a-3p | 0.432623598 | 0.349 |
| hsa-miR-199b-5p | 0.440603314 | 0.223 |
| hsa-miR-3613-3p | 0.44217211 | 0.213 |
| hsa-miR-145-5p | 0.459944222 | 0.463 |
| hsa-miR-597-5p | 0.461633365 | 0.660 |
| hsa-miR-197-5p | 0.466570568 | 0.494 |
| hsa-miR-187-3p | 0.466570568 | 0.396 |
| hsa-miR-324-3p | 0.469786988 | -0.306 |
| hsa-miR-337-3p | 0.469786988 | 0.543 |
| hsa-miR-125a-5p | 0.469786988 | 0.726 |
| hsa-miR-4488 | 0.471536936 | 0.436 |
| hsa-miR-371a-5p | 0.481556544 | 0.137 |
| hsa-miR-942-5p | 0.491690896 | 0.342 |
| hsa-miR-497-5p | 0.499989734 | 0.328 |
| hsa-miR-130b-3p | 0.501938622 | 0.284 |
| hsa-miR-137 | 0.51027761 | 0.344 |
| hsa-miR-769-3p | 0.51027761 | 0.349 |
| hsa-miR-378f | 0.51027761 | 0.463 |
| hsa-miR-4516 | 0.520673198 | -0.186 |
| hsa-miR-24-3p | 0.520673198 | 0.434 |
| hsa-miR-651-5p | 0.520673198 | 0.166 |
| hsa-miR-642a-5p | 0.520673198 | 0.376 |
| hsa-miR-378e | 0.528037945 | -0.051 |
| hsa-miR-548v | 0.533340938 | 0.337 |
| hsa-miR-574-5p | 0.541781177 | 0.440 |
| hsa-miR-542-3p | 0.544033275 | 0.410 |
| hsa-miR-1305 | 0.552490231 | -0.217 |
| hsa-miR-370-3p | 0.552490231 | 0.490 |
| hsa-miR-30b-5p | 0.552490231 | 0.152 |
| hsa-miR-10a-5p | 0.554824729 | 0.182 |
| hsa-miR-576-3p | 0.554824729 | 0.188 |
| hsa-miR-155-5p | 0.563300324 | 0.278 |
| hsa-miR-302a-3p | 0.565719783 | 0.339 |
| hsa-miR-210-3p | 0.571205573 | 0.267 |
| hsa-miR-186-5p | 0.574209619 | 0.412 |
| hsa-miR-551b-3p | 0.574209619 | -0.365 |
| hsa-miR-508-5p | 0.576716552 | 0.404 |
| hsa-miR-1185-2-3p | 0.585216212 | 0.353 |
| hsa-miR-301a-3p | 0.585216212 | 0.455 |
| hsa-miR-525-5p | 0.587813083 | 0.250 |
| hsa-miR-184 | 0.596318135 | -0.237 |
| hsa-miR-4647 | 0.599007348 | 0.291 |
| hsa-miR-570-3p | 0.618799778 | -0.321 |
| hsa-miR-4536-3p | 0.621680635 | 0.213 |
| hsa-miR-454-3p | 0.621680635 | 0.330 |
| hsa-miR-4787-5p | 0.630175244 | 0.198 |
| hsa-miR-4485-3p | 0.63314481 | 0.157 |
| hsa-miR-514a-3p | 0.633155264 | 0.224 |
| hsa-miR-873-3p | 0.641637536 | 0.217 |
| hsa-miR-378h | 0.653184373 | 0.364 |
| hsa-miR-412-3p | 0.656364069 | 0.233 |
| hsa-miR-4536-5p | 0.664813419 | 0.195 |
| hsa-miR-612 | 0.676522279 | 0.572 |
| hsa-miR-603 | 0.676522279 | 0.173 |
| hsa-miR-548ar-5p | 0.676522279 | -0.084 |
| hsa-miR-532-3p | 0.679919372 | 0.224 |
| hsa-miR-937-3p | 0.688308503 | 0.279 |
| hsa-miR-585-3p | 0.697790968 | -0.005 |
| hsa-miR-450a-2-3p | 0.700169686 | -0.022 |
| hsa-miR-3605-5p | 0.703786213 | 0.275 |
| hsa-miR-134-3p | 0.703786213 | 0.203 |
| hsa-miR-136-5p | 0.712102971 | 0.575 |
| hsa-miR-513a-3p | 0.712102971 | 0.226 |
| hsa-miR-95-3p | 0.715827589 | -0.107 |
| hsa-miR-627-3p | 0.71583177 | 0.165 |
| hsa-miR-33b-5p | 0.721881404 | 0.176 |
| hsa-miR-510-3p | 0.72410605 | 0.335 |
| hsa-miR-939-5p | 0.727944464 | 0.216 |
| hsa-miR-1249-3p | 0.727944464 | 0.313 |
| hsa-let-7a-5p | 0.740133578 | 0.124 |
| hsa-miR-639 | 0.760506626 | -0.030 |
| hsa-miR-584-3p | 0.760506626 | 0.162 |
| hsa-miR-1197 | 0.760506626 | 0.078 |
| hsa-miR-203a-5p | 0.764697589 | 0.192 |
| hsa-miR-181d-3p | 0.764697589 | 0.214 |
| hsa-miR-593-3p | 0.764697589 | 0.120 |
| hsa-miR-548ah-5p | 0.772761427 | 0.143 |
| hsa-miR-548a-5p | 0.772761427 | 0.154 |
| hsa-miR-487a-3p | 0.785072171 | 0.041 |
| hsa-miR-411-5p | 0.785072171 | 0.178 |
| hsa-miR-1302 | 0.785072171 | -0.138 |
| hsa-miR-3144-3p | 0.795735984 | 0.099 |
| hsa-miR-127-3p | 0.797435979 | 0.280 |
| hsa-miR-1296-3p | 0.797435979 | -0.083 |
| hsa-miR-3614-5p | 0.801984389 | -0.135 |
| hsa-miR-5010-3p | 0.801984389 | 0.116 |
| hsa-miR-181a-2-3p | 0.809849934 | 0.213 |
| hsa-miR-1323 | 0.809849934 | -0.111 |
| hsa-miR-299-5p | 0.809849934 | 0.117 |
| hsa-miR-767-5p | 0.809849934 | 0.149 |
| hsa-miR-25-5p | 0.814516081 | 0.109 |
| hsa-miR-211-5p | 0.820804197 | 0.114 |
| hsa-miR-29b-3p | 0.822311082 | 0.165 |
| hsa-miR-301b-5p | 0.822311082 | 0.225 |
| hsa-miR-4284 | 0.82710104 | -0.019 |
| hsa-miR-301b-3p | 0.82710104 | 0.197 |
| hsa-miR-502-5p | 0.833409019 | 0.103 |
| hsa-miR-301a-5p | 0.834816434 | 0.185 |
| hsa-miR-1228-3p | 0.839727741 | 0.149 |
| hsa-miR-630 | 0.847362972 | -0.058 |
| hsa-miR-891a-5p | 0.847362972 | -0.088 |
| hsa-miR-1246 | 0.859947643 | 0.111 |
| hsa-miR-365a-3p+hsa-miR-365b-3p | 0.859947643 | 0.282 |
| hsa-miR-200a-3p | 0.87256737 | 0.210 |
| hsa-miR-1253 | 0.877843348 | -0.279 |
| hsa-miR-487b-5p | 0.877843348 | -0.089 |
| hsa-miR-626 | 0.884226072 | -0.109 |
| hsa-miR-512-5p | 0.885219049 | -0.129 |
| hsa-miR-888-5p | 0.890616319 | 0.026 |
| hsa-miR-519c-3p | 0.890616319 | -0.089 |
| hsa-miR-933 | 0.890616319 | 0.256 |
| hsa-miR-627-5p | 0.909828094 | -0.044 |
| hsa-miR-378d | 0.910605731 | 0.152 |
| hsa-miR-514b-5p | 0.923334417 | 0.153 |
| hsa-miR-30e-5p | 0.923334417 | -0.154 |
| hsa-miR-495-5p | 0.923334417 | -0.033 |
| hsa-miR-18b-5p | 0.929091657 | -0.064 |
| hsa-miR-4286 | 0.936082426 | 0.022 |
| hsa-miR-548q | 0.941958787 | -0.065 |
| hsa-miR-563 | 0.941959679 | -0.109 |
| hsa-miR-1255a | 0.94884656 | -0.109 |
| hsa-miR-548j-3p | 0.94884656 | -0.097 |
| hsa-miR-4454+hsa-miR-7975 | 0.961623608 | 0.230 |
| hsa-miR-210-5p | 0.967735685 | -0.070 |
| hsa-miR-183-5p | 0.974186015 | 0.063 |
| hsa-miR-1262 | 0.974186015 | 0.098 |
| hsa-miR-362-5p | 0.980638033 | -0.124 |
| hsa-miR-328-5p | 0.980638033 | 0.153 |
| hsa-miR-302d-3p | 0.987091318 | 0.056 |
| hsa-miR-1185-5p | 0.987203557 | 0.082 |
| hsa-let-7e-5p | 0.987203557 | 0.013 |
| hsa-miR-769-5p | 0.987203557 | -0.024 |
| hsa-miR-1283 | 1 | 0.035 |

| ALL OBSERVATIONS | Cytokine/Chemokine | MW test severe vs mild/moderate | BH adjusted MW test severe vs mild/moderate | MW test mild vs moderate/severe | BH adjusted MW test mild vs moderate/severe |
|---|---|---|---|---|---|
| | GMCSF | 0.100063977 | 0.125079971 | 0.25875553 | 0.457010705 |
| | CCL5 | 0.52267405 | 0.544452135 | 0.005278393 | 0.032989956 |
| | CXCL8 | 0.067398665 | 0.096749488 | 0.153301641 | 0.294810848 |
| | CXCL10 | 0.372562426 | 0.423366393 | 0.09521223 | 0.198358812 |
| | CCL11 | 0.227958563 | 0.271379242 | 0.496041835 | 0.688946993 |
| | CCL17 | 0.099137664 | 0.125079971 | 0.025489533 | 0.07965479 |
| | CCL2 | 0.045302128 | 0.070784575 | 0.08955205 | 0.198358812 |
| | CCL3 | 0.011279619 | 0.021691575 | 0.727617661 | 0.790888762 |
| | CXCL9 | 0.004938951 | 0.010289482 | 0.004582217 | 0.032989956 |
| | CXCL5 | 0.727889221 | 0.727889221 | 0.988333825 | 0.988333825 |
| | CCL20 | 8.14E-05 | 0.000339123 | 0.006777406 | 0.033860481 |
| | CXCL1 | 0.015137621 | 0.025229368 | 0.4250411 | 0.664126718 |
| | CXCL11 | 0.520570998 | 0.544452135 | 0.493987716 | 0.688946993 |
| | CCL4 | 0.069659631 | 0.096749488 | 0.538062623 | 0.707977135 |
| | IL5 | 2.59E-05 | 0.000162123 | 0.968893279 | 0.988333825 |
| | IL2 | 3.38E-05 | 0.000169009 | 0.274206423 | 0.457010705 |
| | IL6 | 2.89E-09 | 7.23E-08 | 1.58E-07 | 3.95E-06 |
| | IL9 | 3.56E-06 | 4.45E-05 | 0.649854563 | 0.757675228 |
| | IL10 | 9.45E-06 | 7.88E-05 | 0.000145574 | 0.001819679 |
| | IFNg | 0.000559963 | 0.001555453 | 0.012348342 | 0.044101223 |
| | TNF | 0.003429262 | 0.007793777 | 0.666754201 | 0.757675228 |
| | IL17A | 9.87E-05 | 0.000352664 | 0.087423684 | 0.198358812 |
| | IL17F | 0.014537906 | 0.025229368 | 0.586254223 | 0.732817779 |
| | IL4 | 0.000127721 | 0.000399127 | 0.008126515 | 0.033860481 |
| | IL22 | 0.001673095 | 0.004182738 | 0.045849743 | 0.127360398 |

| FIRST SAMPLES | Cytokine/Chemokine | MW test severe vs mild/moderate | BH adjusted MW test severe vs mild/moderate | MW test mild vs moderate/severe | BH adjusted MW test mild vs moderate/severe |
|---|---|---|---|---|---|
| | GMCSF | 0.287519621 | 0.399332807 | 0.441750607 | 0.708718903 |
| | CCL5 | 0.227599807 | 0.383365362 | 0.055768895 | 0.464740788 |
| | CXCL8 | 0.355880203 | 0.444850254 | 0.149533586 | 0.55945976 |
| | CXCL10 | 0.008048722 | 0.040243611 | 0.273777224 | 0.570369216 |
| | CCL11 | 0.508921519 | 0.605858951 | 0.1332601 | 0.55945976 |
| | CCL17 | 0.009957223 | 0.041488427 | 0.51027761 | 0.708718903 |
| | CCL2 | 0.108844013 | 0.272110034 | 0.146411685 | 0.55945976 |
| | CCL3 | 0.154809461 | 0.35006839 | 0.259175986 | 0.570369216 |
| | CXCL9 | 0.00286971 | 0.023914252 | 0.18622388 | 0.570369216 |
| | CXCL5 | 0.277209086 | 0.399332807 | 0.607513355 | 0.736666402 |
| | CCL20 | 0.004930121 | 0.030813258 | 0.156648733 | 0.55945976 |
| | CXCL1 | 0.342161278 | 0.444850254 | 0.618799778 | 0.736666402 |
| | CXCL11 | 0.168032827 | 0.35006839 | 0.923334417 | 0.923334417 |
| | CCL4 | 0.599758691 | 0.646127749 | 0.859947643 | 0.923334417 |
| | IL5 | 0.245353832 | 0.383365362 | 0.466536291 | 0.708718903 |
| | IL2 | 0.042930464 | 0.1341577 | 0.250644187 | 0.570369216 |
| | IL6 | 0.000139883 | 0.00349707 | 0.000806266 | 0.020156639 |
| | IL9 | 0.014829605 | 0.052962874 | 0.788587401 | 0.896122047 |
| | IL10 | 0.001243682 | 0.015546022 | 0.00816034 | 0.102004247 |
| | IFNg | 0.70070776 | 0.70070776 | 0.501932045 | 0.708718903 |
| | TNF | 0.544202087 | 0.618411462 | 0.451819218 | 0.708718903 |
| | IL17A | 0.200124212 | 0.383365362 | 0.386712484 | 0.708718903 |
| | IL17F | 0.620282639 | 0.646127749 | 0.922657709 | 0.923334417 |
| | IL4 | 0.100919409 | 0.272110034 | 0.271107039 | 0.570369216 |
| | IL22 | 0.241892884 | 0.383365362 | 0.615944827 | 0.736666402 |

### MILD

| | | | | | | | | | | |
|---|---|---|---|---|---|---|---|---|---|---|
| CCL5 | hsa-miR-1253 | -0.64 | hsa-miR-378e | -0.61 | hsa-miR-1283 | -0.61 | hsa-miR-411-5p | -0.54 | hsa-miR-378h | -0.47 |
| CCL3 | hsa-miR-92a-3p | 0.56 | hsa-miR-1283 | 0.49 | hsa-miR-378e | 0.47 | hsa-miR-340-5p | 0.44 | hsa-miR-106a-5p +hsa-miR-17-5p | 0.43 |
| CXCL9 | hsa-miR-664a-3p | -0.35 | hsa-miR-378d | -0.35 | hsa-miR-28-3p | -0.31 | hsa-miR-548e-5p | -0.29 | hsa-miR-3614-5p | -0.29 |
| CCL20 | hsa-miR-376c-3p | 0.47 | hsa-miR-92a-3p | 0.46 | hsa-miR-382-5p | 0.43 | hsa-miR-378f | -0.39 | hsa-miR-376a-3p | 0.38 |
| CXCL1 | hsa-miR-1283 | -0.52 | hsa-miR-378e | -0.45 | hsa-miR-1253 | -0.45 | hsa-miR-1246 | -0.41 | hsa-miR-873-3p | -0.41 |
| IL5 | hsa-miR-320e | -0.38 | hsa-miR-301b-3p | -0.36 | hsa-miR-1253 | -0.35 | hsa-miR-630 | -0.33 | hsa-miR-378e | -0.32 |
| IL2 | hsa-miR-1255a | -0.41 | hsa-miR-28-3p | -0.4 | hsa-miR-378f | -0.39 | hsa-miR-34a-5p | -0.37 | hsa-miR-301a-5p | -0.36 |
| IL6 | hsa-miR-301a-3p | -0.5 | hsa-miR-26a-5p | -0.46 | hsa-miR-18a-5p | -0.45 | hsa-miR-625-5p | -0.44 | hsa-miR-1305 | -0.43 |
| IL9 | hsa-miR-548j-3p | -0.45 | hsa-miR-122-5p | -0.45 | hsa-miR-1249-3p | -0.43 | hsa-miR-320e | -0.43 | hsa-miR-495-3p | -0.42 |
| IL10 | hsa-miR-513a-3p | -0.45 | hsa-miR-1255a | -0.44 | hsa-miR-1295a | -0.43 | hsa-miR-378f | -0.43 | hsa-miR-664a-3p | -0.42 |
| IFNg | hsa-miR-378e | -0.5 | hsa-miR-548q | -0.48 | hsa-miR-764 | -0.47 | hsa-miR-644a | -0.47 | hsa-miR-1253 | -0.47 |
| TNF | hsa-miR-376c-3p | 0.38 | hsa-miR-518b | 0.35 | hsa-miR-495-3p | 0.34 | hsa-miR-128-3p | 0.32 | hsa-miR-152-3p | 0.32 |
| IL17A | hsa-miR-4707-5p | -0.36 | hsa-miR-128-3p | 0.36 | hsa-miR-548g-3p | -0.32 | hsa-miR-301a-5p | -0.32 | hsa-miR-376c-3p | 0.31 |
| IL17F | hsa-miR-92a-3p | 0.47 | hsa-miR-548g-3p | -0.34 | hsa-miR-1255a | -0.34 | hsa-miR-320e | 0.34 | hsa-miR-221-3p | 0.32 |
| IL4 | hsa-miR-1255a | -0.4 | hsa-miR-301a-5p | -0.36 | hsa-miR-513a-3p | -0.36 | hsa-miR-4707-5p | -0.35 | hsa-miR-433-3p | 0.34 |
| IL22 | hsa-miR-548ah-5p | -0.31 | hsa-miR-1255a | -0.28 | hsa-miR-155-5p | -0.27 | hsa-miR-92a-3p | 0.27 | hsa-miR-1197 | -0.25 |
| CXCL10 | hsa-miR-1283 | -0.3 | hsa-miR-582-5p | -0.28 | hsa-miR-26a-5p | -0.27 | hsa-miR-28-3p | -0.24 | hsa-miR-378e | -0.24 |
| CCL17 | hsa-miR-92a-3p | 0.53 | hsa-miR-361-5p | 0.5 | hsa-miR-423-5p | 0.44 | hsa-miR-143-3p | 0.43 | hsa-miR-221-3p | 0.42 |

### MODERATE

| | | | | | | | | | | |
|---|---|---|---|---|---|---|---|---|---|---|
| CCL5 | hsa-miR-19a-3p | 0.43 | hsa-miR-342-3p | 0.43 | hsa-miR-221-3p | 0.41 | hsa-miR-27b-3p | 0.41 | hsa-miR-15a-5p | 0.4 |
| CCL3 | hsa-miR-365a-3p +hsa-miR-365b-3p | 0.34 | hsa-miR-421 | 0.33 | hsa-miR-574-3p | 0.31 | hsa-miR-551b-3p | 0.3 | hsa-miR-1283 | 0.28 |
| CXCL9 | hsa-miR-223-3p | -0.46 | hsa-miR-3613-3p | 0.46 | hsa-miR-181a-3p | 0.43 | hsa-miR-525-5p | 0.43 | hsa-miR-25-5p | 0.42 |
| CCL20 | hsa-miR-223-3p | -0.57 | hsa-miR-126-3p | -0.46 | hsa-miR-495-3p | -0.43 | hsa-miR-199a-3p +hsa-miR-199b-3p | -0.42 | hsa-miR-23a-3p | -0.42 |
| CXCL1 | hsa-miR-1283 | -0.39 | hsa-miR-451a | -0.34 | hsa-miR-144-3p | -0.32 | hsa-miR-4454 +hsa-miR-7975 | 0.3 | hsa-let-7a-5p | 0.3 |
| IL5 | hsa-miR-133a-3p | -0.32 | hsa-miR-187-3p | -0.29 | hsa-miR-30d-5p | -0.28 | hsa-miR-576-3p | -0.28 | hsa-miR-376c-3p | 0.25 |
| IL2 | hsa-miR-30d-5p | -0.29 | hsa-miR-221-3p | -0.28 | hsa-miR-15a-5p | -0.27 | hsa-miR-335-5p | -0.26 | hsa-miR-340-5p | -0.26 |
| IL6 | hsa-miR-320e | 0.45 | hsa-miR-223-3p | -0.39 | hsa-miR-495-3p | -0.39 | hsa-miR-630 | 0.37 | hsa-miR-191-5p | -0.35 |
| IL9 | hsa-miR-510-3p | -0.43 | hsa-miR-626 | -0.42 | hsa-miR-576-3p | -0.39 | hsa-miR-10b-5p | -0.39 | hsa-miR-642a-5p | -0.37 |
| IL10 | hsa-miR-342-3p | -0.3 | hsa-miR-4454 +hsa-miR-7975 | 0.3 | hsa-miR-29c-3p | -0.26 | hsa-miR-320e | 0.26 | hsa-miR-29a-3p | -0.25 |
| IFNg | hsa-miR-451a | 0.41 | hsa-miR-378h | -0.36 | hsa-miR-888-5p | -0.35 | hsa-miR-510-3p | -0.34 | hsa-miR-642a-5p | -0.33 |
| TNF | hsa-miR-3168 | -0.33 | hsa-miR-133a-3p | -0.29 | hsa-miR-335-5p | -0.29 | hsa-miR-576-3p | -0.28 | hsa-miR-451a | 0.28 |
| IL17A | hsa-miR-510-3p | -0.25 | hsa-miR-28-5p | -0.24 | hsa-miR-187-3p | -0.24 | hsa-miR-612 | -0.24 | hsa-miR-548ah-5p | -0.24 |
| IL17F | hsa-miR-548ah-5p | -0.32 | hsa-miR-133a-3p | -0.29 | hsa-miR-1268b | -0.28 | hsa-miR-337-5p | -0.28 | hsa-miR-1283 | 0.26 |
| IL4 | hsa-let-7d-5p | 0.31 | hsa-miR-371a-5p | 0.31 | hsa-miR-1305 | 0.27 | hsa-miR-125b-5p | 0.25 | hsa-miR-377-3p | 0.24 |
| IL22 | hsa-miR-223-3p | -0.54 | hsa-miR-191-5p | -0.49 | hsa-miR-503-5p | -0.42 | hsa-miR-23a-3p | -0.41 | hsa-miR-29b-3p | -0.41 |
| CXCL10 | hsa-miR-191-5p | -0.5 | hsa-miR-150-5p | -0.47 | hsa-miR-106a-5p +hsa-miR-17-5p | -0.47 | hsa-miR-223-3p | -0.44 | hsa-miR-93-5p | -0.44 |
| CCL17 | hsa-miR-365a-3p +hsa-miR-365b-3p | 0.49 | hsa-miR-421 | 0.47 | hsa-miR-335-5p | 0.4 | hsa-miR-425-5p | 0.37 | hsa-miR-181a-2-3p | 0.36 |

### SEVERE

| | | | | | | | | | | |
|---|---|---|---|---|---|---|---|---|---|---|
| CCL5 | hsa-miR-1537-3p | 0.48 | hsa-miR-340-5p | 0.46 | hsa-miR-148b-3p | 0.46 | hsa-miR-299-3p | 0.45 | hsa-miR-197-3p | 0.45 |
| CCL3 | hsa-miR-30d-5p | -0.54 | hsa-miR-425-5p | -0.52 | hsa-miR-19a-3p | -0.52 | hsa-miR-16-5p | -0.51 | hsa-miR-19b-3p | -0.5 |
| CXCL9 | hsa-miR-1972 | 0.35 | hsa-miR-6721-5p | 0.26 | hsa-miR-630 | 0.25 | hsa-miR-570-3p | 0.24 | hsa-miR-1285-5p | 0.22 |
| CCL20 | hsa-miR-342-3p | -0.46 | hsa-miR-425-5p | -0.45 | hsa-miR-19b-3p | -0.43 | hsa-miR-19a-3p | -0.43 | hsa-miR-376c-3p | 0.43 |
| CXCL1 | hsa-miR-630 | 0.32 | hsa-miR-4516 | 0.3 | hsa-miR-212-3p | -0.23 | hsa-miR-873-3p | -0.31 | hsa-miR-122-5p | -0.17 |
| IL5 | hsa-miR-1246 | -0.48 | hsa-miR-551b-3p | -0.47 | hsa-miR-30d-5p | -0.47 | hsa-miR-16-5p | -0.45 | hsa-miR-25-3p | -0.44 |
| IL2 | hsa-miR-425-5p | -0.53 | hsa-miR-19a-3p | -0.51 | hsa-miR-30d-5p | -0.51 | hsa-miR-19b-3p | -0.47 | hsa-miR-222-3p | -0.46 |
| IL6 | hsa-miR-19b-3p | -0.54 | hsa-miR-342-3p | -0.48 | hsa-miR-19a-3p | -0.48 | hsa-miR-425-5p | -0.46 | hsa-miR-30d-5p | -0.46 |
| IL9 | hsa-miR-374b-5p | -0.45 | hsa-miR-127-3p | -0.42 | hsa-miR-1246 | -0.41 | hsa-miR-136-5p | -0.41 | hsa-miR-30b-5p | -0.4 |
| IL10 | hsa-miR-630 | 0.4 | hsa-miR-19b-3p | -0.39 | hsa-miR-19a-3p | -0.38 | hsa-miR-518b | -0.36 | hsa-miR-503-5p | -0.33 |
| IFNg | hsa-miR-425-5p | -0.58 | hsa-miR-16-5p | -0.57 | hsa-miR-19b-3p | -0.56 | hsa-miR-30d-5p | -0.56 | hsa-miR-19a-3p | -0.54 |
| TNF | hsa-miR-425-5p | -0.51 | hsa-miR-30d-5p | -0.5 | hsa-miR-222-3p | -0.48 | hsa-miR-376c-3p | -0.48 | hsa-miR-24-3p | -0.48 |
| IL17A | hsa-miR-769-3p | -0.39 | hsa-miR-1246 | -0.39 | hsa-miR-551b-3p | -0.38 | hsa-miR-181d-3p | -0.36 | hsa-miR-1973 | -0.35 |
| IL17F | hsa-miR-30d-5p | -0.51 | hsa-miR-425-5p | -0.49 | hsa-miR-376c-3p | -0.47 | hsa-miR-495-3p | -0.47 | hsa-miR-19a-3p | -0.46 |
| IL4 | hsa-miR-30d-5p | -0.57 | hsa-miR-425-5p | -0.57 | hsa-miR-24-3p | -0.54 | hsa-miR-29a-3p | -0.54 | hsa-miR-19a-3p | -0.54 |
| IL22 | hsa-miR-451a | -0.4 | hsa-miR-485-3p | -0.39 | hsa-miR-136-5p | -0.36 | hsa-miR-376c-3p | -0.36 | hsa-miR-382-5p | -0.35 |
| CXCL10 | hsa-miR-1285-5p | 0.63 | hsa-miR-1972 | 0.58 | hsa-miR-574-5p | 0.54 | hsa-miR-4454 +hsa-miR-7975 | 0.53 | hsa-miR-1246 | 0.53 |
| CCL17 | hsa-miR-1246 | -0.48 | hsa-miR-7-5p | -0.42 | hsa-miR-302d-3p | -0.41 | hsa-miR-1285-5p | -0.4 | hsa-miR-127-3p | -0.4 |

| | | | | | | | | | | |
|---|---|---|---|---|---|---|---|---|---|---|
| hsa-miR-24-3p | IL4 | -0.54 | IFNγ | -0.53 | CCL3 | -0.5 | TNF | -0.48 | IL2 | -0.46 |
| hsa-let-7f-5p | CXCL1 | 0.34 | CCL5 | 0.29 | IL10 | -0.15 | IL17A | 0.11 | CXCL5 | 0.1 |
| hsa-miR-630 | IL9 | 0.4 | IFNγ | 0.4 | CCL17 | 0.33 | IL5 | 0.32 | CCL3 | 0.28 |
| hsa-miR-29b-3p | IL4 | -0.45 | IL2 | -0.41 | CCL3 | -0.4 | GMCSF | -0.38 | IFNγ | -0.37 |
| hsa-let-7a-5p | CCL5 | 0.32 | CXCL1 | 0.28 | CXCL10 | 0.28 | IL4 | -0.25 | CCL20 | -0.25 |
